# Interplay Between the Gut Microbiome and Typhoid Fever: Insights from Endemic Countries and a Controlled Human Infection Model

**DOI:** 10.1101/2024.09.02.24312347

**Authors:** Philip M. Ashton, Leonardos Mageiros, James E. Meiring, Angeziwa Chunga Chirambo, Farhana Khanam, Sabina Dongol, Happy Banda, Abhilasha Karkey, Lorena Preciado Llanes, Helena Thomaides-Brears, Malick Gibani, Nazmul Hasan Rajib, Nazia Rahman, Prasanta Kumar Biswas, Md Amirul Islam Bhuiyan, Sally Kay, Kate Auger, Olivier Seret, Nicholas R. Thomson, Andrew J Pollard, Stephen Baker, Buddha Basnyat, John D. Clemens, Christiane Dolecek, Sarah J. Dunstan, Gordon Dougan, Robert S. Heyderman, Virginia E. Pitzer, Firdausi Qadri, Melita A. Gordon, Kathryn E. Holt, Thomas C. Darton, STRATAA Study Group

## Abstract

Typhoid fever is a systemic infection caused by *Salmonella enterica* serovar Typhi (*S.* Typhi) invasion from the gut lumen. Transmission between people occurs through ingestion of contaminated food and water, particularly in settings with poor water and sanitation infrastructure, resulting in over 10 million illnesses annually. As the pathogen invades via the gastrointestinal tract, it is plausible that the gut microbiome may influence the outcome of *S*. Typhi exposure. There is some evidence that bacteria producing short-chain fatty acids (SCFAs) may create an environment unfavourable to invasive *Salmonella,* but data from humans is limited.

To investigate the association between gut microbiome and typhoid fever, we analysed samples collected from three all-age cohorts enrolled in a prospective surveillance study conducted across three settings where typhoid fever is endemic (Dhaka, Bangladesh; Blantyre, Malawi; and Kathmandu, Nepal). Cohorts consisted of acute typhoid fever patients (n=92), asymptomatic household contacts of typhoid fever patients (representing individuals who were likely exposed to *S*. Typhi but did not develop disease, n=97), and asymptomatic serosurvey participants with high Vi antibody titres (representing individuals who were exposed to *S*. Typhi and may be carriers, n=69). The stool microbiomes of each cohort were characterised using shotgun metagenomics, and bacterial diversity, composition, and function were compared.

We identified 4 bacterial species that were significantly lower in abundance in typhoid fever patients compared with household contacts (i.e. probably exposed), in two of the three participant populations (Bangladesh and Malawi). These bacteria may represent taxa that provide protection against development of clinical infection upon exposure to *S*. Typhi, and include the inflammation-associated species *Prevotella copri* clade A and *Haemophilus parainfluenzae*. Our functional analysis identified 28 specific metabolic gene clusters (MGCs) negatively associated with typhoid fever in Bangladesh and Malawi, including seven MGCs involved in SCFA metabolism. The putative protection provided by microbiome SCFA metabolism was supported by data from a controlled human infection model conducted in a UK population, in which participants who did not develop typhoid fever following ingestion of *S*. Typhi had higher abundance of a putative SCFA-metabolising MGC (q-value = 0.22).

This study identified the same protective associations between taxonomic and functional microbiota characteristics and non-susceptibility to typhoid fever across multiple human populations. Future research should explore the potential functional role of SCFAs and inflammation-associated bacteria in resistance to *S.* Typhi and other enteric infections.

## Introduction

Typhoid fever, caused by invasive *Salmonella enterica* serovar Typhi (*S.* Typhi) infection, causes an estimated 10.9 million illnesses and 116,800 deaths per year disproportionately affecting people in South and Southeast Asia and sub-Saharan Africa (Stanaway et al. 2019; Meiring et al. 2023). The introduction of typhoid conjugate vaccines is expected to decrease the number of typhoid fever cases in these settings, along with improvements in water, sanitation and hygiene (WASH). However, the large burden of disease, the role of asymptomatic gall bladder carriage as a source of infection, and the fact that typhoid fever often impacts the most impoverished and marginalised communities, means that eradicating this infection will be difficult (Meiring et al. 2021; Patel et al. 2021; Steele et al. 2016).

The human gut microbiota, a complex ecosystem comprising trillions of microbial cells, plays an indispensable role in shaping our overall health and susceptibility to diseases (Shreiner, Kao, and Young 2015). This densely populated microbial environment is not only pivotal in the processes of digestion and nutrient absorption but also intimately connected to our immune system, determining its responses to various stimuli (Belkaid and Hand 2014). As the nexus between the external environment and our internal physiology, the gut microbiome is a critical determinant of the outcome of gastrointestinal infectious disease exposures, via a phenomenon termed ‘colonization resistance’ (Stecher et al. 2007; Winter et al. 2010; Lopez et al. 2016; Libertucci and Young 2019). This protective effect arises from multifaceted interactions: direct competition for nutrients, production of antimicrobial compounds, and modulation of the host’s innate and adaptive immune responses (Leshem, Liwinski, and Elinav 2020). Some of the earliest work on colonisation resistance showed that suppressing the gut microbiome of mice with antibiotics dramatically reduced the dose of *Salmonella* required to establish infection (Bohnhoff, Drake, and Miller 1954). Recent work has made great strides in understanding mechanisms by which *Salmonella* and the microbiome interact (Rogers, Tsolis, and Bäumler 2021). Short chain fatty acids (SCFAs) are important in interaction between the microbiota and *Salmonella*, reducing the availability of oxygen in the gut (Byndloss et al. 2017) and acidifying the cytosol of *Salmonella,* inhibiting growth (Jacobson et al. 2018). Despite well-established knowledge about the gut microbiome’s function in preventing *Salmonella* infections and the established faecal-oral transmission pathway of *S*. Typhi, the precise influence of the gut microbiome on the outcome of *S*. Typhi exposure remains underexplored, complicated by the human host-specificity of *S*. Typhi.

While insights from animal studies of invasive infection with other *S. enterica* serovars can be informative, detailed studies of the interaction between the human gut microbiome and *S.* Typhi specifically are lacking. Controlled human infection model (CHIM) studies provide an experimental system in which to interrogate infections in humans, complementing observational studies of natural infections (Waddington et al. 2014; Gibani et al. 2020). Previous data generated from this model revealed an association between typhoid fever susceptibility and the presence of *Methanobrevibacter* in the gut microbiome (Zhang et al. 2018). Analysing natural infections, Haak et al. determined that typhoid fever patients in Bangladesh had reduced microbial diversity and fewer SCFA producers than healthy counterparts (Haak et al. 2020). Better understanding of the causal relationships between the gut microbiome and typhoid fever could lead to novel preventative mechanisms and diagnostics, as well as an improved understanding of colonisation resistance to bacteria causing serious invasive diseases in high burden settings.

To fill this knowledge gap, we sequenced the stool microbiome of 258 participants enrolled in the STRATAA study, conducted in three diverse high typhoid fever burden settings in Asia and Africa (Meiring et al. 2021). Signals that replicated in more than one study population were further investigated in a UK-based CHIM cohort.

## Materials & Methods

### Participants and sample collection

Detailed methods for the STRATAA study have been published previously (Darton et al. 2017; Meiring et al. 2021). In brief, approximately 100,000 individuals were enumerated in a demographic census in three communities: Ndirande in Blantyre, Malawi; Lalitpur in Kathmandu, Nepal; and Mirpur in Dhaka, Bangladesh. Febrile patients, of any age, within the study populations were recruited via passive surveillance, and “acute typhoid fever” patients were defined as those with positive blood cultures yielding *S.* Typhi. When participants presented with fever, stool samples were collected where possible, preferably prior to antimicrobial use. Stool specimens were transferred to -80°C within 6 hours of collection. Data on antimicrobial use prior to their enrolment was recorded (Darton et al. 2017). Blood-culture confirmed typhoid fever patients were then followed up and stool samples were collected from their household contacts; asymptomatic stool culture-negative individuals were included in microbiome sequencing. High Vi-titre individuals were identified from a community-based serological survey of up to 8,500 age-stratified participants per site from the original demographic census. From the serological survey, samples were analysed for anti-Vi IgG antibody (Meiring et al. 2021). The participants at each site with the highest Vi responses were followed up and stool sample collected for microbiome analysis (Khanam et al. 2021). Stool cultures were *S*. Typhi-negative in all but one participant (Khanam et al. 2021), however it is known that carriers shed only intermittently and high-Vi individuals is an acceptable predictor for *S.* Typhi carriage (Robbins and Robbins 1984; Qureshi et al. 2024). For the microbiome sub-study, we randomly selected for sequencing stool samples from up to 40 participants from each site, from each of three groups: acute typhoid fever cases, household contacts, and serosurvey participants with high-Vi titres.

Research ethics committee approval for a joint study protocol across all three surveillance sites was obtained within each country and from the Oxford Tropical Research Ethics Committee (University of Oxford, Oxford, UK).

### DNA extraction and sequencing

DNA extraction from stool was done using the Fast-Prep 24 FastDNA Spin Kit (MP Biomedicals, CA, USA) according to manufacturer’s instructions. DNA was sent to the Wellcome Sanger Institute (UK) for metagenomic sequencing using Illumina HiSeq 2500 or HiSeq 4000 to generate 150 bp paired end reads, yielding an average 14.8 million read pairs (standard deviation 2.3 million) per sample.

### Bioinformatics

Raw sequencing data was quality trimmed and adapters were removed using bbduk v38.96 with the parameters ‘ktrim=r k=23 mink=11 hdist=1 tbo tpe qtrim=r trimq=20 minlength=50’. Sequence deriving from human DNA was removed by mapping to a human reference genome (GCF_009914755.1) and the hostile tool (Constantinides, Hunt, and Crook 2023). Taxonomic profiling was carried out with metaphlan v4.0.6 with database version mpa_vOct22_CHOCOPhlAnSGB_202212 (Blanco-Míguez et al. 2023).

We used the BiG-MAP program (Victória Pascal Andreu et al. 2021) (https://github.com/medema-group/BiG-MAP, commit e7b8042) to compare metagenomic readsets against a database of non-redundant metabolic gene clusters (MGCs) identified using gutSMASH (Victòria Pascal Andreu et al. 2023) from a collection of reference genomes from the Culturable Genome Reference, the Human Microbiome Project and other Clostridia genomes (https://zenodo.org/records/7252625#.ZFVTrexBz0r). Each MGC is assigned to a species which is the species from which the representative sequence representing the cluster was obtained. BiG-MAP calculated the number of reads per kilobase per million reads (RPKM) for each metabolic gene cluster.

A sub-sample of 8.8 million reads was taken from each readset using the ‘samplè command of the seqtk tool (v1.3-r106). Sub-sampled reads were then analysed with Resistance Gene Identifier (RGI) v6.0.2 bwt command against the CARD v3.2.7 reference database (Alcock et al. 2023). Due to uncertainty regarding some CARD database classifications (e.g. OXA-1 was classified as a carbapenemase, a curation error which has subsequently been resolved), we used the drug class information for each gene from the NCBI AMR Reference Gene Database (refgene) v2023-08-08.2 (https://www.ncbi.nlm.nih.gov/pathogens/refgene). Only “Core” genes from NCBI refgene were analysed; these genes are almost entirely mobilisable AMR determinants and do not include mutational resistance. Extended-spectrum beta-lactamases (ESBLs) were identified based on the gene product descriptions in NCBI refgene. Only genes with an Average Percent Coverage of 100% in the RGI analysis were included in further analysis. The number of reads mapped to each gene (RGI Completely Mapped Reads) was normalised by the length of the gene (RGI Reference Length) to generate a “Reads per kilobase of AMR gene” metric. There was no need to normalise for the number of reads, as all readsets had been sub-sampled to the same number of reads. RGI results were parsed and analysed using the scripts described in the Data Availability section of this manuscript. AMR genes identified in *S.* Typhi were obtained from (Argimón et al. 2021).

### Statistical analysis

Statistical analyses were done in R (v4.1.0). To quantify alpha diversity, Shannon’s index was calculated using the alpha.div function in the rbiom package (v1.0.3), and ANOVA analysis carried out with the aov function (stats package, v4.1.0). The Bray-Curtis metric of beta diversity was calculated with the vegdist function of the R package vegan (v2.6.4). Principal co-ordinates analysis (PCoA) was carried out with the cmdscale function (package stats, v4.1.0), and statistics comparing beta diversity between participant groups (i.e. acute typhoid, household contacts, presumptive carrier) was carried out using a PERMANOVA approach implemented by the adonis2 function of the vegan package (v2.6.4). Associations between taxa and phenotypes of interest (e.g. participant group) were explored using multivariable linear modelling implemented in the maaslin2 function of the maaslin2 R package (v1.6.0) (Mallick et al. 2021). Depending on the analysis, co-variates such as age, sex, antibiotic exposure, and sequencing run were included, as described. For maaslin2 analyses, Benjamini-Hochberg correction was applied for multiple testing. For analyses of data from endemic cohorts a q-value threshold of 0.05 was used to identify significant associations, while for CHIM data analyses, due to small sample size, the default maaslin2 q-value threshold of 0.25 was used.

### Controlled human infection model analysis

We used data from a *Salmonella* Typhi and Paratyphi A CHIM cohort to validate the associations identified in the endemic countries. Briefly, healthy adults aged 18-60 were recruited to be challenged with a single oral dose of either *S.* Typhi or *S.* Paratyphi A (Gibani et al. 2020). The primary endpoint for the model was a diagnosis of typhoid or paratyphoid fever, defined as a temperature of ≥38°C persisting for ≥12 hours and/or *S*. Typhi/Paratyphi A bacteraemia in a sample collected ≥72 hours after oral challenge. Only data from participants who were being challenged for the first time were analysed here. Stool samples were taken from participants prior to challenge and stored at -80°C. DNA extraction, sequencing and analyses was undertaken using the methods described above. The primary maaslin2 analyses for this cohort included only species or MGC classes that were significantly associated with participant group in at least two endemic country cohorts; a secondary analysis was carried out without this restriction. Analyses were done separately for participants challenged with either *S.* Typhi or *S.* Paratyphi A, in addition to a combined analysis including both pathogens.

## Results

### Description of population

Stool microbiome sequences were successfully generated for 258 participants from Bangladesh (n = 80), Malawi (n = 102), and Nepal (n = 76). Across the three populations, there were three participant groups; 92 patients with acute typhoid fever, 97 household contacts of acute typhoid cases, and 69 people with high anti-Vi titres. The baseline characteristics of participants are described in Table 1. Overall, acute typhoid fever patients were significantly younger than household contacts, while in Bangladesh and Malawi high-Vi titre participants were significantly older than household contacts (Supplementary Figure 1, Table 1). Overall, in Malawi, Bangladesh, and Nepal, 62%, 53% and 54% of participants were female, respectively (Table 1, Supplementary Figure 2). In Bangladesh and Malawi, sex distribution was similar between the participant groups, whereas in Nepal, 82% of household contacts and 67% of carriers were female, while only 24% of typhoid fever cases were female (Table 1, Supplementary Figure 2). Of the acute typhoid fever cases, 37.5%, 73.9% and 44.8% from Bangladesh, Malawi, and Nepal, respectively, reported antimicrobial use in the 2 weeks prior to stool sample collection.

**Table 1:** Characteristics of participants in the microbiome sub-study of STRATAA surveillance.

|  |  | **Bangladesh** | **Malawi** | **Nepal** |
| --- | --- | --- | --- | --- |
| **Acute typhoid** | N | 40 | 23 | 29 |
|  | Median age, years (range) | 6 (1-60) | 10 (3-40) | 17 (5-38) |
|  | Female (%) | 48% | 61% | 24% |
|  | Antibiotics in last 2 weeks (%) | 38% | 74% | 45% |
| **Carrier** | N | NA* | 39 | 30 |
|  | Median age | NA* | 32 | 43.9 |
|  | Female (%) | NA* | 56% | 67% |
|  | Antibiotics in last 2 weeks (%) | NA* | NA† | NA† |
| **Control** | N | 40 | 40 | 17 |
|  | Median age, years (range) | 29 (4-65) | 24 (18-41) | 35 (15-48) |
|  | Female (%) | 65% | 65% | 82% |
|  | Antibiotics in last 2 weeks (%) | NA† | NA† | NA† |

\* Carriers from Bangladesh were not included in the analysis due to sample protocol differences
† Antibiotic usage in the two weeks previous was an exclusion criterion for carriers and household contacts.

### Acute typhoid fever patients compared with household contacts

First, we compared stool metagenomic profiles of acute typhoid patients with those of household contacts, within each of the three study populations, to explore microbiome signatures associated with typhoid fever. Participant group was not significantly associated with differences in alpha diversity in any population (Supplementary Figure 3, Supplementary Table 1). Beta diversity varied significantly between typhoid fever patients and household contacts across all sites (Figure 1, Supplementary Table 2). The proportion of beta diversity variance explained by participant group was highest in Malawi (R^2^ = 0.25, FDR = 0.01), followed by Bangladesh (R^2^ = 0.07, FDR = 0.01), Nepal (R^2^ = 0.06, FDR = 0.01 and the combined analysis (R^2^ = 0.04, FDR = 0.01) (Figure 1, Supplementary Table 2, Supplementary Table 3, Supplementary Table 4, Supplementary Table 5). Participant group explained the greatest beta diversity variance of the factors investigated (which also included sex, age, and reported antibiotic use) in a combined analysis of all three sites and for each site individually. In Malawi, prior antibiotic usage and the interaction of prior antibiotic usage with age and sex were also significantly associated with beta diversity, although these variables explained a lower proportion of variance than participant group (R^2^ 0.02-0.05, Supplementary Table 4). In Nepal, sex was significantly associated with beta diversity and explained almost as much variance as participant group (R^2^ = 0.04, Supplementary Table 5), which was itself associated with sex at this site. Firmicutes was the dominant phylum in typhoid fever patients and household contacts at all three sites, followed by Bacteroidetes in Malawi and Nepal and Actinobacteria in Bangladesh (Supplementary Figure 4, Supplementary Table 6).

**Figure 1:**
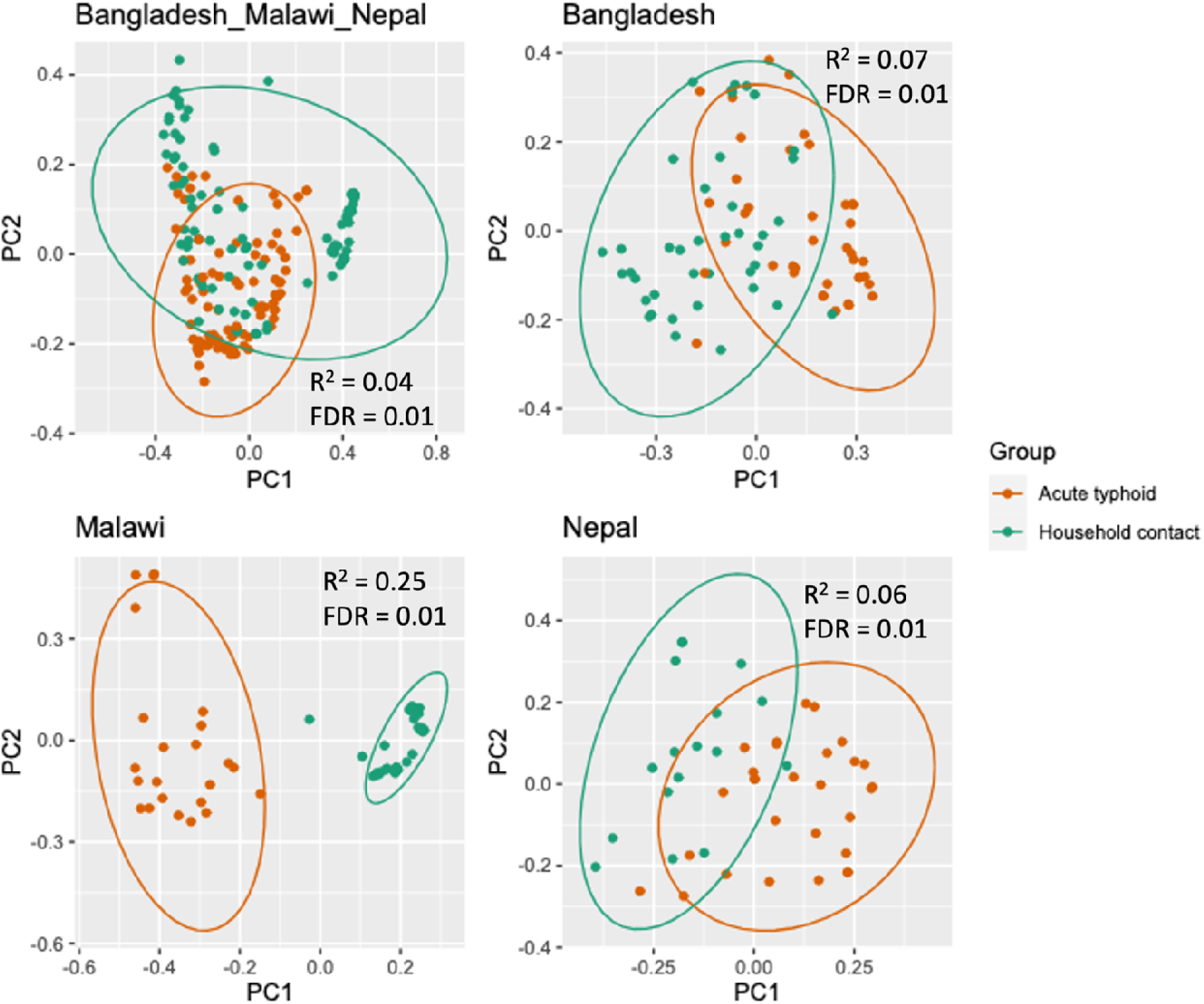
Principal co-ordinate analysis of the beta diversity between household contacts and typhoid fever participants across all three sites, and for each site individually. The proportion of variance explained (R^2^) and the Bonferroni corrected FDR from a PERMANOVA analysis including participant group, sex, and antibiotic usage are displayed on each plot. In the combined analysis, country of sampling was also included in the PERMANOVA analysis.

Maaslin2 analysis of taxonomic profiles identified 92, 23 and 0 species significantly associated with household contact vs typhoid fever participant groups in Malawi, Bangladesh, and Nepal, respectively (Figure 2, Supplementary Table 7, Supplementary Table 8). No taxa showed significant associations across all three sites; however, four showed consistent associations at two sites (Bangladesh and Malawi), all of which were negatively associated with typhoid fever (Figure 2). These four species were *Prevotella copri* clade A, *Haemophilus parainfluenzae, Clostridium* SGB6179 and a *Veillonellaceae* spp represented by a metagenome assembled genome, GGB4266_SGB5809. Although there were differences in age between cases and household contacts at all sites, age was included as a covariate in maaslin2, and the associations with typhoid status were also evident within age groups where data from that age group was available (Supplementary Figures 5-8). Of these four species, none were significantly associated with typhoid fever in Nepal or the CHIM; most samples from the CHIM did not contain any reads assigned to these species (Supplementary Table 29).

**Figure 2:**
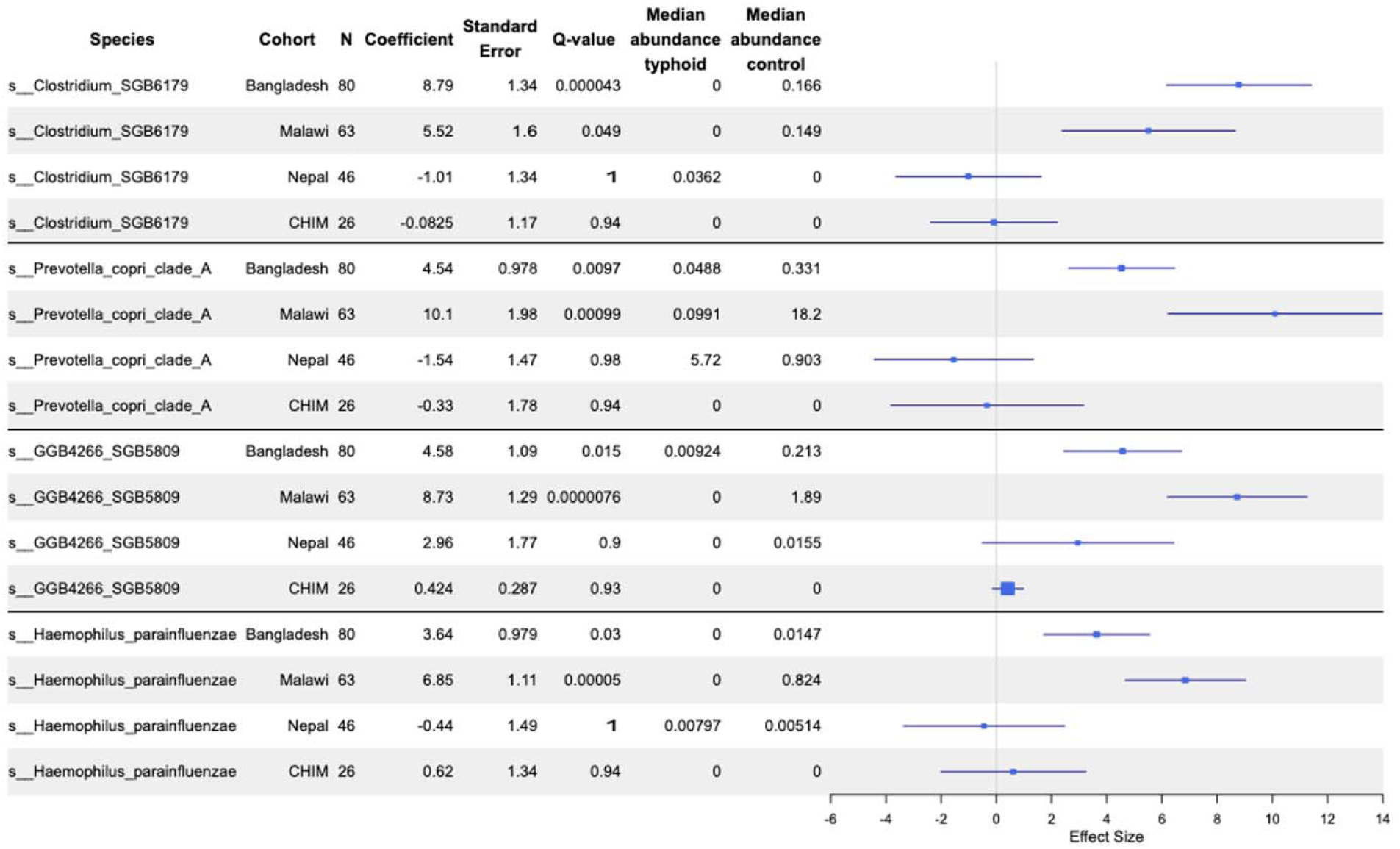
Forest plot of species that significantly differed between household contacts and typhoid fever participants in at least two endemic countries. Results from all three endemic countries and the CHIM cohort are shown for context. CHIM maaslin2 analysis only included species that were significantly associated in at least two of the endemic country sites. Species are labelled with their GTDB identifier, note that ‘s GGB4266_SGB5809’ refers to a proposed novel genus/species of Veillonellaceae.

Some species were associated with typhoid fever at a single endemic country site. In Bangladesh, there were 19 species for which the relative abundance significantly differed between typhoid fever participants and household contacts, that didn’t differ at any other site, or in the CHIM (Supplementary Table 7). Three species were negatively associated with typhoid fever (*Prevotella copri* clade C, *Romboutsia timonensis*, and *Ligilactobacillus* ruminis), while 16 species including 7 species of Actinomyces were positively associated with typhoid fever (Supplementary Table 7). In Malawi, 70 species were negatively associated with typhoid fever, including *Ruminococcus gnavus*, *Roseburia intestinalis*, *Roseburia inulinivorans,* and *Faecalibacterium prausnitzii* (Supplementary Table 8), while 18 species were positively associated with typhoid fever including 11 (61%) that are only in the Metaphlan4 database as metagenome assembled genomes (Supplementary Table 8).

To complement our taxonomic analysis, we also carried out functional gene analysis. We explored two dimensions of variation in metabolic gene clusters (MGCs). First, we examined the distribution of specific MGCs, investigating participant group differences at a granular, sub-type level (e.g. a specific RNF complex from a strain of *Bacteroides ovatus*). Second, we compared the abundance of different MGC types between participant groups (e.g. all RNF complex MGCs). This dual approach enables us to capture both detailed and broad patterns of functional gene distribution within the microbiome.

We identified 264 specific MGCs that significantly differed between typhoid fever patients and household contacts in the Malawi cohort, 126 in Bangladesh, and 2 in Nepal (Supplementary Table 24, Supplementary Table 25, Supplementary Table 26). Neither of the specific MGC associations identified in the Nepal data were replicated in the Malawi or Bangladesh cohorts. There were 28 specific MGCs significantly associated with typhoid fever in both Bangladesh and Malawi (summarised in Table 2, full information in Supplementary Table 9), all were negatively associated with typhoid fever in both settings. Six of these specific MGCs were linked to SCFA metabolism (‘pyruvate2acetate.formate’) and five to anaerobic metabolism (‘Rnf complex’), associated with *Prevotella* and *Haemophilus* (see Figure 2, Supplementary Table 9). None of these 28 specific MGCs showed negative associations with acute typhoid fever in Nepal.

**Table 2:** Summary of the specific MGCs negatively associated with typhoid fever in both Bangladesh and Malawi, and the species of each MGC reference sequence. Full information in Supplementary Table 9.

| MGC_class | n | Species |
| --- | --- | --- |
| Pyruvate2acetate.formate | 6 | Haemophilus aegyptius, Haemophilus haemolyticus, Haemophilus parainfluenzae, Prevotella copri, Prevotella sp., Prevotella sp. |
| Rnf_complex | 5 | Haemophilus parainfluenzae, Prevotella copri, Prevotella sp., Prevotella sp., Prevotella sp. |
| TPP_AA_metabolism | 4 | Clostridium celatum, Haemophilus pittmaniae, Haemophilus sputorum, Prevotella sp. |
| Respiratory_glycerol | 2 | Aggregatibacter sp., Haemophilus parainfluenzae |
| Arginine2putrescine.Putrescine2spermidine | 1 | Romboutsia sp. |
| Formate_dehydrogenase | 1 | Haemophilus sp. |
| Fumarate2succinate | 1 | Haemophilus sp. |
| OD_eut_pdu_related.PFOR_II_pathway | 1 | Paeniclostridium sordellii |
| OD_fatty_acids | 1 | Dialister succinatiphilus |
| Others_HGD_unassigned | 1 | Prevotella copri |
| PFOR_II_pathway | 1 | Prevotella sp. |
| TPP_AA_metabolism.Arginine2putrescine | 1 | Haemophilus sp. |
| TPP_fatty_acids | 1 | Prevotella copri |
| porA | 1 | Prevotella copri |
| succinate2propionate | 1 | Dialister invisus |

There were 31 MGC types that significantly differed between typhoid fever patients and household contacts in the Malawi cohort, 8 in Bangladesh, and 1 in Nepal (Figure 3, Supplementary Table 30). There was one MGC type that was negatively associated with typhoid fever in both Malawi and Bangladesh, “TPP_AA_metabolism.Arginine2putrescine”. Other MGC types that were negatively associated with typhoid fever in Malawi only included “acetate2butyrate.TPP_fatty_acids”, “TPP_fatty_acids.aminobutyrate2Butyrate”, “acrylate2propionate” and “Rnf_complex.succinate2propionate”, while in Bangladesh, “Pyruvate2acetate” was negatively associated with typhoid fever. In Bangladesh, “Others_HGD_unassigned.Nitrate_reductase” was positively associated with typhoid fever, while in Malawi, two MGC types were positively associated with typhoid fever – “Molybdopterin_dependent_oxidoreductase” and “pdu”. In Nepal, “Fumarate2succinate.fatty_acids” was significantly negatively associated with typhoid fever.

**Figure 3:**
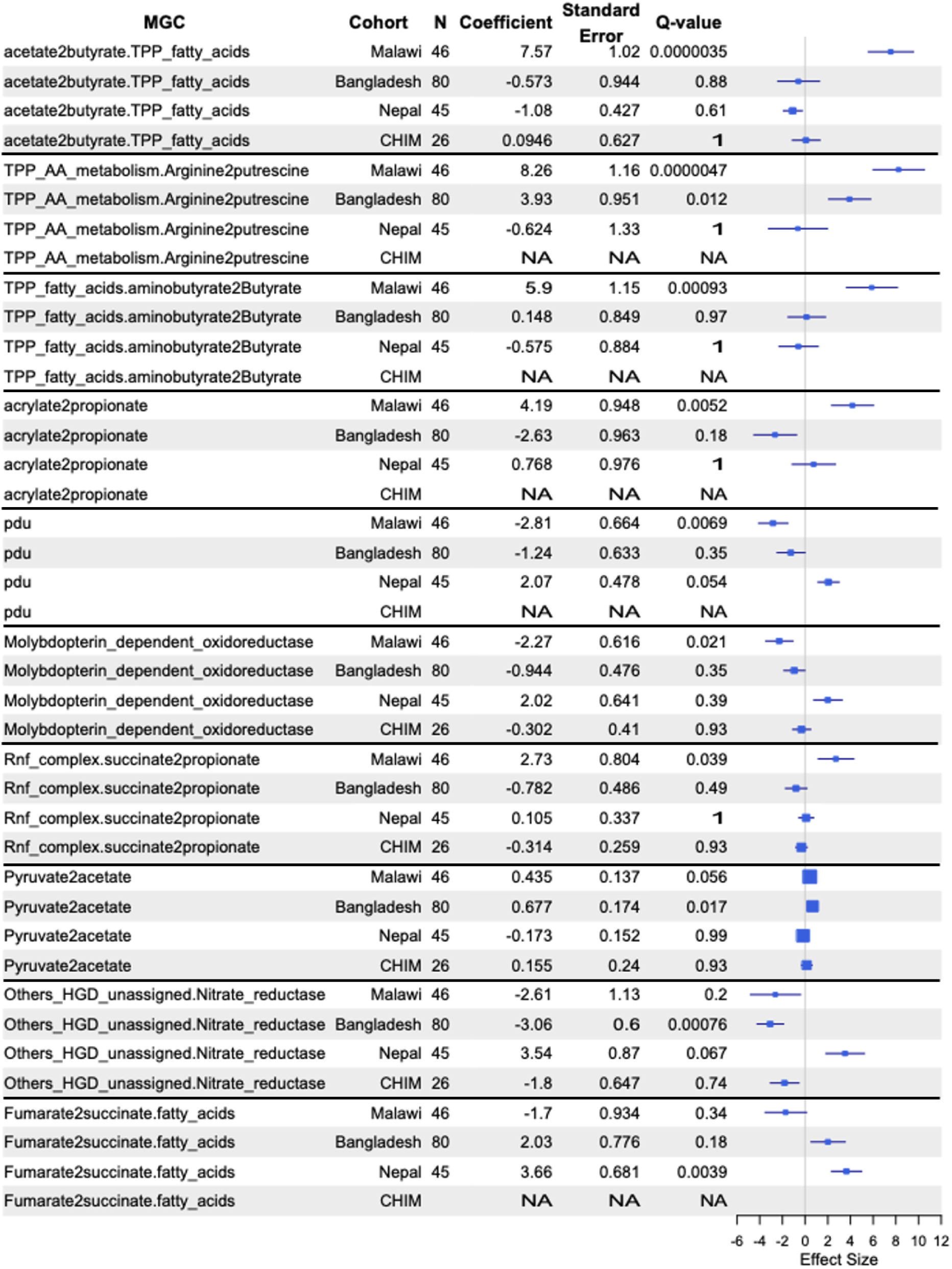
Forest plot of selected MGC types that differed significantly between household contacts and typhoid fever participants in at least one cohort. If an entry is NA, then there was insufficient of that MGC type identified in that cohort for maaslin analysis.

We hypothesised the taxa and MGCs that were found in higher abundance in asymptomatic household contacts of typhoid fever patients may be protective against developing disease upon exposure to *S*. Typhi. We sought to explore this more directly by assessing their abundance in CHIM participants who did or did not develop enteric fever upon challenge with typhoidal *Salmonella*, in a UK-based study (Gibani et al. 2020). We sequenced the microbiomes of 13 participants who were subsequently challenged with *S.* Typhi and 13 with *S.* Paratyphi A, of which 7 and 6 were diagnosed with enteric (typhoid or paratyphoid) fever, respectively (Supplementary Table 18). There was no significant difference (FDR < 0.05) in alpha or beta diversity at baseline among CHIM participants by age at challenge, sex, or subsequent enteric fever diagnosis (Supplementary Table 19, Supplementary Table 20, Supplementary Figure 15). In an analysis including only the 28 MGC classes that were identified as negatively associated with typhoid in both Malawi and Bangladesh, two were also negatively associated with typhoid fever diagnosis in CHIM participants: an “Rnf_complex.Glycine_cleavage.succinate2propionate” MGC (coefficient = -3.4, q-value = 0.22) and an “Rnf_complex” MGC (coefficient = -6.2, q-value = 0.22) (Supplementary Figure 16, full results available in Supplementary Table 23). None of the 28 MGC classes tested were associated with paratyphoid fever or a combination of both typhoid and paratyphoid fever. We investigated whether any of the species associated with typhoid in both Malawi and Bangladesh were associated with susceptibility to typhoid and/or paratyphoid fever in the CHIM cohort; none of them were (see Figure 2). All four species that were associated with typhoid fever in the endemic cohorts were not detected in the majority of samples from the CHIM (Supplementary Table 29).

### Analysis of high-Vi participants

High-Vi participants may represent asymptomatic gall-bladder carriers, or individuals with recent exposure or sub-clinical infection, either of whom may be a source in transmission. Detection of carriers is important for typhoid fever control, and will remain so even as disease incidence reduces due to vaccination and improvements in WASH, therefore we sought to explore the microbiome signature of this group. Samples from Bangladesh were not included in this analysis, as laboratory processing of the high Vi-titre participant samples from Bangladesh was not consistent with the other sites or participant groups. Using PERMANOVA, participant group was significantly associated with beta diversity in both countries (age and sex were not significant, Supplementary Table 11, Supplementary Table 12). Participant group explained greater variance in the Malawi cohort (R^2^ 0.25, FDR=0.01) than Nepal (R^2^ 0.07, FDR=0.01, Supplementary Table 11, Supplementary Table 12, Supplementary Figure 10). The three participant groups have distinct beta-diversity signatures, with high-Vi individuals closer to typhoid patients than to household contacts (Figure 4). In the Malawi cohort, 125 bacterial species significantly differed between household contacts and high Vi-titre individuals (Supplementary Table 13), and 41 of these also significantly differed between household contacts and typhoid fever (Supplementary Table 21). These 41 species include *H. parainfluenzae, Blautia obeum,* and *Ruminococcus gnavus* (all less prevalent in acute typhoid and high-Vi individuals than household contacts, Supplementary Figure 11). Amongst Nepal samples, only one species significantly differed between household controls and high-Vi individuals (the Firmicutes SGB GGB9790 SGB15413). There were no species associated with household contacts compared with typhoid patients from Nepal, and SGB15413 did not differ between these groups (Supplementary Figure 12).

**Figure 4:**
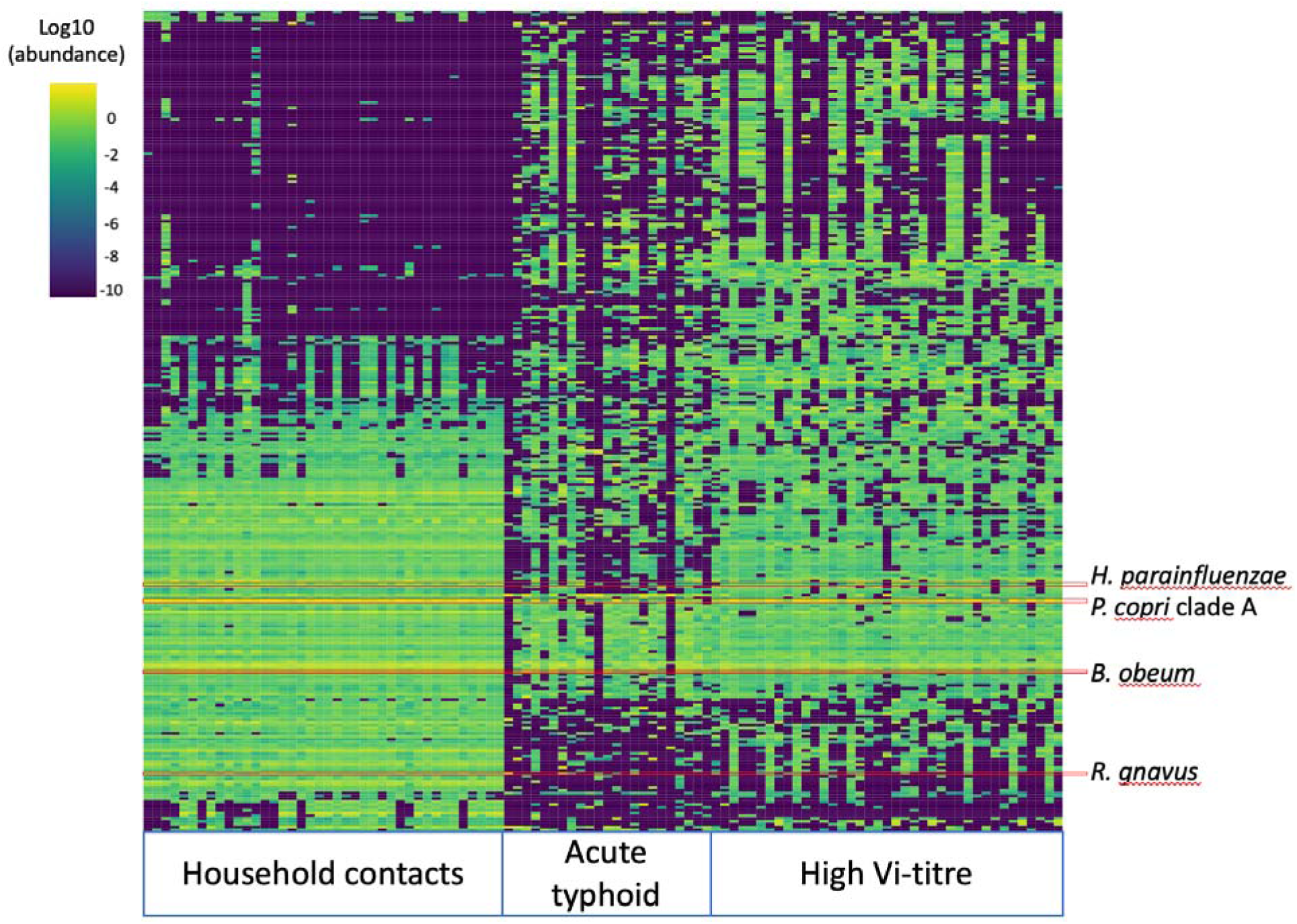
Heatmap of the abundance of the 400 most abundant species in household contacts, acute typhoid fever cases and high Vi-titre participants from Malawi. The columns are sorted by participant group, rows are sorted by hierarchical clustering across all samples.

The combined relative abundance of species only described as species genome bins (SGBs) was significantly lower in household contacts in Malawi (13.9%), compared with both typhoid fever patients (34.8%, P=0.012) and high Vi-titre participants (47.1%, P=2.2×10^−16^) in this setting (Supplementary Figure 13). The 50 SGBs with the highest summed abundance across all participant types in Malawi belonged to Firmicutes (n = 41), Actinobacteria (n = 4), Bacteroidetes (n = 3), and Proteobacteria (n = 2) (Supplementary Table 22). The most common families of the 50 most abundant SGBs were Oscillospiraceae (n = 13), Lachnospiraceae (n = 7), and Clostridiaeceae (n = 4) (Supplementary Table 22). In Nepal, high Vi-titre individuals had lower SGB relative abundance than acute typhoid cases (16.8% vs 29.9%, p = 0.03, Supplementary Figure 13).

### AMR genes

We recently reported AMR genotypes and phenotypes for *S*. Typhi isolated from acute typhoid fever cases in Bangladesh, Malawi, and Nepal (Dyson et al. 2024). Quinolone resistance was common in Bangladesh and Nepal but rare in Malawi; here we saw a similar pattern reflected in the gut microbiomes, with 2% quinolone resistance in Malawi, 30% in Bangladesh and 15% in Nepal (only mobile genetic element encoded quinolone determinants were analysed in the metagenomic data, Supplementary Table 14). Macrolide resistance was identified in one *S*. Typhi isolate from Bangladesh (0.6%) but not in Nepal or Malawi, here we found higher rates of macrolide resistance in microbiomes from individuals in Bangladesh (38%) and Nepal (35%) and lower in Malawi (11%). For the older drugs, *S*. Typhi isolates from Malawi had near-universal resistance to older sulfonamides and tetracyclines, and this was mirrored in high rates of resistance genes to these drugs in the microbiome samples (92% sulfonamides, 52% tetracycline). ESBLs were not detected in any *S*. Typhi isolates but were common in the microbiomes in Nepal (17%) and Bangladesh (9%); less so in Malawi (3%).

The proportion of samples with macrolide and trimethoprim resistance genes that have been observed in *S.* Typhi was significantly higher in participants with acute typhoid fever than in household contacts or high Vi-titre participants (macrolides = 42%, 14%, 22% respectively, Chi-square test p-value =4.4×10^−5^; trimethoprim = 32%, 12%, 20% respectively, Chi-square test p-value = 0.005). There was no significant difference between participant types in the proportion of samples with resistance genes to any other drug classes. There was no statistically significant association between prior reported antibiotic usage and the number of AMR genes (Wilcoxon-rank sum P-value = 0.52) or the number of AMR gene classes (Wilcoxon Rank Sum P-value = 0.56) in typhoid fever patients across all sites (Supplementary Figure 14).

## Discussion

In this study, we examined the relationship between the stool microbiome and typhoid fever in three settings with endemic disease. Our analyses identified four microbial species (Figure 2), linked to anaerobic fermentation and SCFA metabolism (Table 2), that differentiated typhoid fever patients from household contacts in Bangladesh and Malawi. While these taxa were not found in the UK-based CHIM participants, a different MGC with a potential link to SCFA metabolism was higher in participants who were not susceptible to typhoid fever. Looking at the abundance of different MGC types (as opposed to specific MGCs) we found four types of MGC involved in SCFA metabolism that were negatively associated with typhoid fever in Malawi, and one in Bangladesh. Overall, our data support a protective role for SCFA-producing microbes, although with our study design it is not possible to deconvolute the impact of the microbiome on typhoid from the impact of typhoid on the microbiome.

The strengths of our study include a larger sample size than previous investigations of the interaction between typhoid fever and the microbiome, and a multi-site study design to increase generalisability. In contrast to previous studies of the microbiome and typhoid fever, we used shotgun metagenomics to characterise the gut flora, enabling in depth functional analysis and investigation of AMR genes. Notably, using the same methodology to analyse participants from different countries, most of the taxa we associated with typhoid fever differed between locations. This suggests that susceptibility to disease in different human populations may be modified by different species, which could hamper the generalisability of study findings between populations. It is notable that the most consistent microbial signature for disease susceptibility, identified across multiple study populations with small sample sizes, was a functional signature (specifically gene clusters associated with SCFA metabolism) rather than any specific taxa that perform this function.

One of the key limitations of our study is the lack of age and sex matching between the typhoid fever and household contact groups. The age and sex composition of the household controls reflects the occupants of the household during the daytime visits of the study workers. This discrepancy may introduce bias, as age and sex are known to influence the composition and function of the microbiome (Yatsunenko et al. 2012). Although these variables were included in the statistical analyses, the lack of matching weakens our findings. Our use of a case-control study design in the endemic countries generated numerous hypotheses for subsequent research, but it does not prove causal relationships. Additionally, we did not collect dietary information from participants. Diet is a major determinant of microbiome composition and function, and variations in dietary habits could confound the associations we observed (Bourdeau-Julien et al. 2023). Lastly, accounting for the effect of antibiotics on the microbiome in these cohorts presents a complex challenge, as it can significantly alter microbiome composition and potentially mask or mimic associations with typhoid fever. These challenges could largely be addressed using CHIM studies. Unfortunately, the only available CHIM data was from a different study population (in the UK) and with a small sample size (n=13 exposed to *S*. Typhi). Use of larger high-income country CHIMs and development of CHIMs in populations from settings with endemic disease would be helpful to better elucidate specific microbial signatures of protection.

Our use of shotgun metagenomics enabled us to also investigate the prevalence of AMR genes associated with resistance in *S.* Typhi across the three sites and participant groups. Macrolide and trimethoprim resistance genes were more common in acute typhoid cases compared with the household controls and high Vi-titre participants, highlighting that typhoid and associated antimicrobial usage imposes a selective pressure on the gut microbiome. This supports the idea that reducing disease via immunisation could reduce AMR beyond just *S.* Typhi. Higher levels of resistance to sulphonamides in Malawi likely reflects the common use of trimethoprim-sulfamethoxazole in HIV/AIDS programs (MacPherson et al. 2022), while higher microbiome prevalence of acquired ESBL and fluoroquinolone-resistance genes in Bangladesh than Malawi reflects the local epidemiology (e.g. recent studies of blood stream infections show that in Bangladesh, 72% and 75% were resistant to ciprofloxacin and third-generation cephalosporins respectively, compared with 31% and 30% in Malawi) (Musicha et al. 2017; Ahmed et al. 2017). The gut microbiome prevalence of resistance genes followed a similar pattern to the prevalence of resistance genes in *S.* Typhi observed from these sites (Dyson et al. 2024) for tetracycline, sulphonamides and quinolones, which likely reflects a shared evolutionary pressure on the isolates and the microbiome. The fact that non-ESBL beta-lactamases were identified in fewer metagenomes than ESBL beta-lactamases is because we only counted beta-lactamase encoding genes that are commonly found in *S.* Typhi.

Zhang et al. investigated 16S community profile and metatranscriptome features associated with susceptibility to typhoid fever in a CHIM (Zhang et al. 2018). One of their primary findings was that the archaeal genus *Methanobrevibacter* was enriched in people who were challenged with *S.* Typhi but did not develop disease. In contrast, we only identified one association with a *Methanobrevibacter* species (*M. smithii*); a positive association with typhoid fever in Malawi. Furthermore, Zhang et al., identified that *Prevotella* was higher in people who developed typhoid fever, in contrast to our findings that it was negatively associated with typhoid fever in natural infections in endemic settings. One source of these discrepancies could be that Zhang et al. employed 16S, while we used shotgun metagenomics. Haak et al. observed more SCFA producing bacteria in, in both healthy control and febrile but non-typhoid fever participants in their study in Bangladesh compared with typhoid fever patients, and higher faecal load of SCFAs in controls than any febrile group (Haak et al. 2020), which corresponds well with our findings. It has also been shown that higher SCFAs in an acidic environment, and a strict anaerobic environment, can inhibit the growth of Enterobacteriaceae such as *Salmonella* via both direct and indirect mechanisms (Byndloss et al. 2017; Jacobson et al. 2018; Rogers, Tsolis, and Bäumler 2021). Recently, higher relative abundance of butyrate producing bacteria was associated with a reduced risk of hospitalisation for infections in a prospective study carried out in high income European countries (Kullberg et al. 2024). Two MGC types positively associated with typhoid fever in Malawi were molybdopterin dependent oxidoreductase and propanediol utilisation microcompartments, both of which enable growth in the inflamed gut (Faber et al. 2017; Zhu et al. 2018), suggesting that the microbiota of typhoid fever participants in Malawi may be in a state of dysbiosis. Arginine to putrescine was the only MGC type associated with health in both Bangaldesh and Malawi, polyamines such as putrescine promote gut barrier integrity (Rao, Xiao, and Wang 2020), and can act as immunomodulators (Proietti et al. 2020).

Our findings from the STRATAA cohorts can be interpreted from two perspectives: i) the microbiota protecting against typhoid and, ii) typhoid causing microbiota changes. In contrast, the CHIM study design enables us to specifically assess whether microbiota present prior to pathogen exposure are associated with the outcome of that exposure (i.e. developing disease or not). Unfortunately, none of the taxa negatively associated with typhoid fever in the endemic settings were present in most of the CHIM participants, making it impossible to validate the association. It is well established that microbiome species composition differs between human populations, and particularly between those in high-income countries and low-and-middle-income countries (Gupta, Paul, and Dutta 2017). However, the metabolic gene cluster findings identified a recurring finding across the endemic settings and the CHIM; an association between species encoding SCFA metabolising genes and non-susceptibility to typhoid fever. While it should be noted that the CHIM sample size was small, the fact that we identified a statistically significant difference supports the idea that the effect is quite strong. While our CHIM analyses lacked power, which is reflected in the high q-values obtained, the q-value for the SCFA MGC association was below the default maaslin2 threshold for significant findings.

Both *P. copri* and *H. parainfluenzae,* which were significantly lower in typhoid fever patients compared with household contacts in both Bangladesh and Malawi, are associated with increased gut inflammation (Sohn et al. 2023; Larsen 2017). *P. copri* is thought to increase Th17 inflammation (Godinez et al. 2011) while *H. parainfluenzae* stimulates intestinal (IFN-γ)+ CD4+ T cells (Sohn et al. 2023), two mechanisms which play a key role in the response to *Salmonella* infections (Bao et al. 2000; Raffatellu et al. 2008). Increased inflammation could prime the host to respond more rapidly to pathogenic exposures, leading to enhanced control of infections. It is highly plausible that the gut microbiome plays a role in shaping the response of the immune system to pathogen challenge (Durack and Lynch 2019). This intriguing association needs further investigation, as the role of inflammation in enabling *Salmonella* to overcome colonisation resistance in mice is well established (Winter et al. 2010; Rogers, Tsolis, and Bäumler 2021). Among the species negatively associated with typhoid fever in Malawi only was *Ruminococcus gnavus*, which could influence susceptibility to enteric infection via IgA stimulation (Bunker et al. 2019), protects against enteropathogenic *E. coli* (McGrath et al. 2022), stimulates host tryptophan catabolism (Hoffmann et al. 2016) (*S.* Typhi requires tryptophan to grow in macrophages (Blohmke et al. 2016)), and produces secondary bile acids including chenodeoxycholic acid and iso-LCA that have anti-virulence effects on *Salmonella* (Lee et al. 2013; Yang, Stein, and Hang 2023).

Direction of causality cannot be determined from our study design, and a murine model of *Salmonella* Typhimurium infection recently reported shifts in gut microbiome composition, including a reduction in *Ruminococcaceae* taxa associated with acetate and butyrate production, following infection (Rogers et al. 2024). It is therefore plausible that the microbiome is modified in people suffering from typhoid fever. Gut disruption is known to alter the gut microbiome, for example, colorectal cancer patients from Morocco and Kenya had reduced *P. copri* in their gut, (Allali et al. 2018; Obuya et al. 2022). Potential triggers for microbiome alterations in typhoid fever could include antibiotic exposure, dietary changes or anorexia due to sickness, *S*. Typhi-induced malabsorption in the small intestine resulting in alterations to the nutrient composition within the large intestine, and the direct impact of fever on microbiome composition as the increase in temperature could favour the growth of particular bacterial species (Chuttani, Jain, and Misra 1971; Huus and Ley 2021).

We cannot be certain about the exact cause of higher levels of anti-Vi antibodies in the high Vi-titre participants. Potential sources of immune stimulation include recent asymptomatic *S.* Typhi exposure/infection, exposure/infection with *Citrobacter freundii* which can be Vi-antigen positive, or chronic *S.* Typhi carriage. It is striking that there was a clear divergence in microbiota profile between these participants and acute cases and household contacts in Malawi, as this suggests that there is an interaction between the cause of the high Vi-titre and the gut microbiota. The 41 species that were significantly lower in both typhoid fever cases and high Vi-titre participants included the potentially immune-modulatory *H. parainfluenzae* (Sohn et al. 2023), the bile acid modifying *B. obeum* (formerly *Ruminococcus obeum*), and *R. gnavus*. Notably, it has been demonstrated that bile salt hydrolases encoded by *B. obeum* can inhibit *Vibrio cholerae* virulence gene activation and colonization (Alavi et al. 2020), and it is intriguing to hypothesise a similar mechanism might protect against *S*. Typhi gallbladder carriage and/or systemic infection. The influence of these species on susceptibility to *S.* Typhi infection, gallbladder carriage, and immune-reactivity should be investigated further.

## Conclusions

We found that typhoid fever patients in Malawi and Bangladesh display distinct microbiome signatures compared to household contacts, marked by lower abundance of SCFA producers and inflammation-related species. The negative association between SCFA producing genes and typhoid fever susceptibility was also replicated to some extent in our UK-based challenge study. The differences in species composition in the microbiota of these distinct cohorts make direct comparison and therefore validation difficult, highlighting the importance of establishing challenge studies in endemic settings to directly address important mechanistic questions.

## Data availability

The code used in the analysis of these data is available on github - https://github.com/flashton2003/STRATAA_metagenomics. Sequencing data for this project are available at the European Nucleotide Archive, project accessions PRJEB14050 and PRJEB22175.

## Acknowledgements

We thank all the STRATAA participants for their willingness to take part in this research. We also acknowledge the sequencing and pathogen informatics teams at the Wellcome Sanger Institute for sequencing and data processing, and Zoe Dyson for assisting with sequence data transfer. We would like to acknowledge Ndaru Jambo, Andreas Bäumler, and Renee Tsolis for helpful discussions. KA, SK, NRT, OS were funded by the Wellcome Trust (220540/Z/20/A). For the purpose of open access, the authors have applied a CC-BY public copyright licence to any author-accepted manuscript version arising from this submission. This research was funded by the Wellcome Trust (STRATAA grant number 106158/Z/14/Z, and core funding to the Wellcome Sanger Institute, grant number 206194) and the Bill & Melinda Gates Foundation (grant number OPP1141321). MAG, ACC, and PMA were supported by a research professorship (NIHR300039) from the National Institute for Health and Care Research, UK Department of Health and Social Care. GD is supported by the Health@InnoHK, Innovation Technology Commission Funding. MMG is supported by the NIHR Imperial Biomedical Research Centre and the Wellcome Trust (224029/Z/21/Z).

Members of the STRATAA study group are Christoph Blohmke, Yama Farooq, Jennifer Hill, Nhu Tran Hoang, Tikhala Makhaza Jere, Harrison Msuku, Tran Vu Thieu Nga, Rose Nkhata, Sadia Isfat Ara Rahman, Nazia Rahman, Neil J Saad, Trinh Van Tan, Deus Thindwa, Merryn Voysey, and Richard Wachepa. Their affiliations are: Malawi-Liverpool-Wellcome Programme, Blantyre, Malawi (Tikhala Makhaza Jere, Harrison Msuku, Rose Nkhata, Deus Thindwa, Richard Wachepa); International Centre for Diarrhoeal Disease Research, Dhaka, Bangladesh (Sadia Isfat Ara Rahman, Nazia Rahman); Oxford Vaccine Group, Department of Paediatrics, University of Oxford, and the NIHR Oxford Biomedical Research Centre, Oxford, UK (Christoph Blohmke, Yama Farooq, Jennifer Hill, Merryn Voysey); The Hospital for Tropical Diseases, Wellcome Trust Major Overseas Programme, Oxford University Clinical Research Unit, Ho Chi Minh City, Viet Nam (Nhu Tran Hoang, Tran Vu Thieu Nga, Trinh Van Tan); Department of Epidemiology of Microbial Diseases and the Public Health Modeling Unit, Yale School of Public Health, Yale University, New Haven, CT, USA (Neil J Saad)

## Author contributions

Philip M. Ashton – conceptualisation, data curation, formal analysis, investigation, methodology, resources, software, visualisation, writing – original draft preparation.

Leonardos Mageiros - conceptualisation, data curation, formal analysis, investigation, methodology, software, writing – review and editing.

James Meiring - data curation, investigation, methodology, writing – review and editing.

Angeziwa Chunga Chirambo – data curation, investigation, writing – review and editing.

Farhana Khanam – data curation, investigation, writing – review and editing.

Happy Banda – investigation, writing – review and editing.

Abhilasha Karkey – investigation, supervision, writing – review and editing.

Sabina Dongol – data curation, investigation, writing – review and editing.

Lorena Preciado Llanes – conceptualisation, data curation, investigation, methodology, writing – review and editing.

Helena Thomaides-Brears - investigation, methodology, writing – review and editing.

Malick Gibani – data curation, investigation, methodology, writing – review and editing.

Nazmul Hasan Rajib – investigation, writing – review and editing.

Nazia Rahman – investigation, writing – review and editing.

Prasanta Kumar Biswas – investigation, writing – review and editing.

Md Amirul Islam Bhuiyan – investigation, writing – review and editing.

Andrew Pollard – conceptualisation, funding acquisition, supervision, writing – review and editing.

Stephen Baker – funding acquisition, writing – review and editing.

Buddha Basnyat – funding acquisition, supervision, resources, writing – review and editing.

John D Clemens – funding acquisition, supervision, writing – review and editing.

Christiane Dolecek – funding acquisition, writing – review and editing.

Sarah J Dunstan – funding acquisition, writing – review and editing.

Gordon Dougan – conceptualisation, funding acquisition, methods, resources, writing – review and editing.

Robert S. Heyderman – funding acquisition, writing – review and editing.

Virginia E. Pitzer – funding acquisition, writing – review and editing.

Firdausi Qadri – funding acquisition, supervision, resources, writing – review and editing.

Melita A. Gordon – conceptualisation, funding acquisition, resources, supervision, writing – review and editing.

Thomas C. Darton – conceptualisation, methodology, writing – review and editing.

Kathryn E. Holt – conceptualisation, funding acquisition, resources, methodology, supervision, writing – original draft preparation, writing – review and editing.

## Supplementary Figures

**Supplementary Figure 1:**
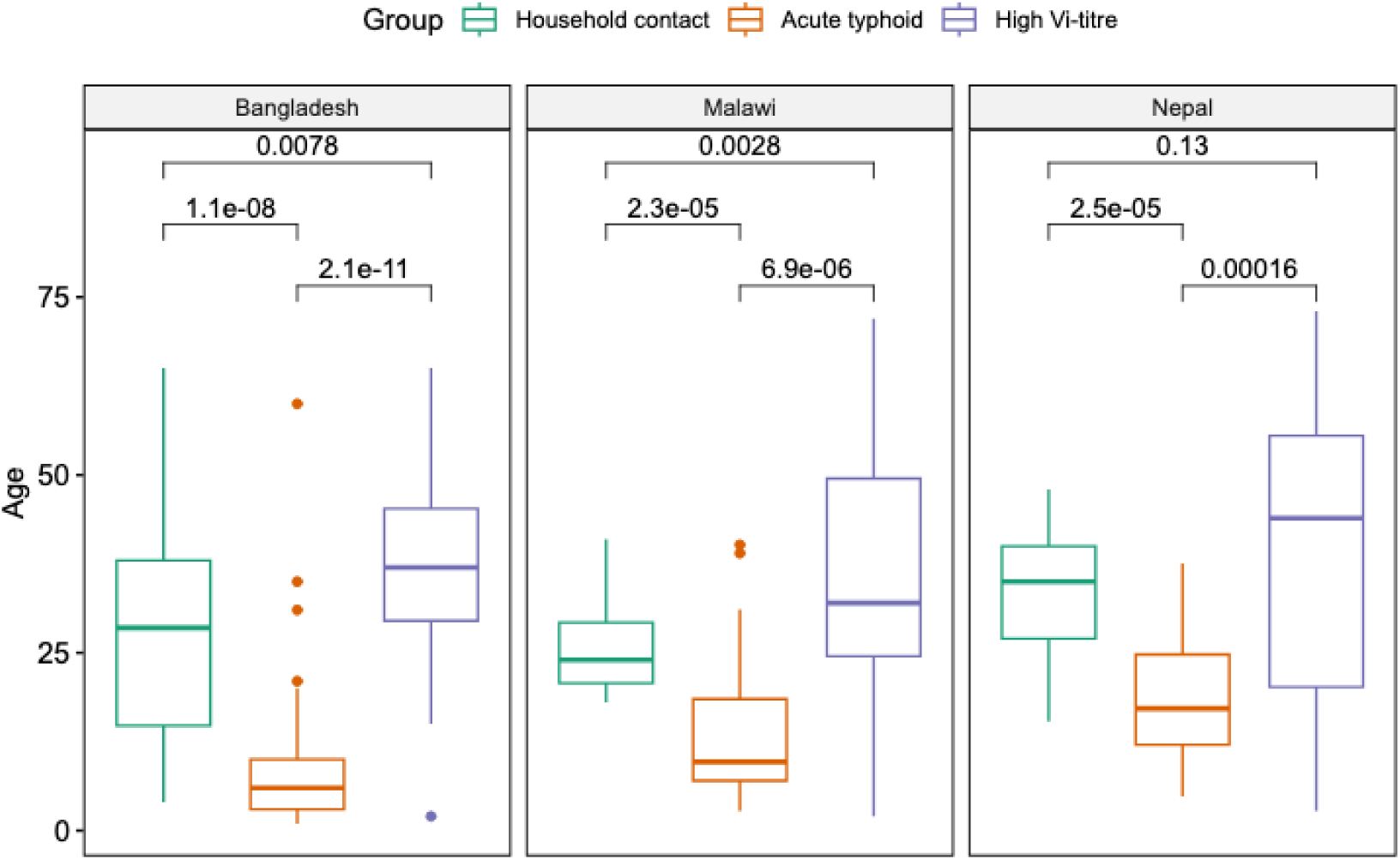
Age distribution of participants in the healthy control, acute typhoid, and presumptive carrier groups, broken down by study site. Statistical annotations are the p-values resulting from Wilcoxon rank sum tests.

**Supplementary Figure 2:**
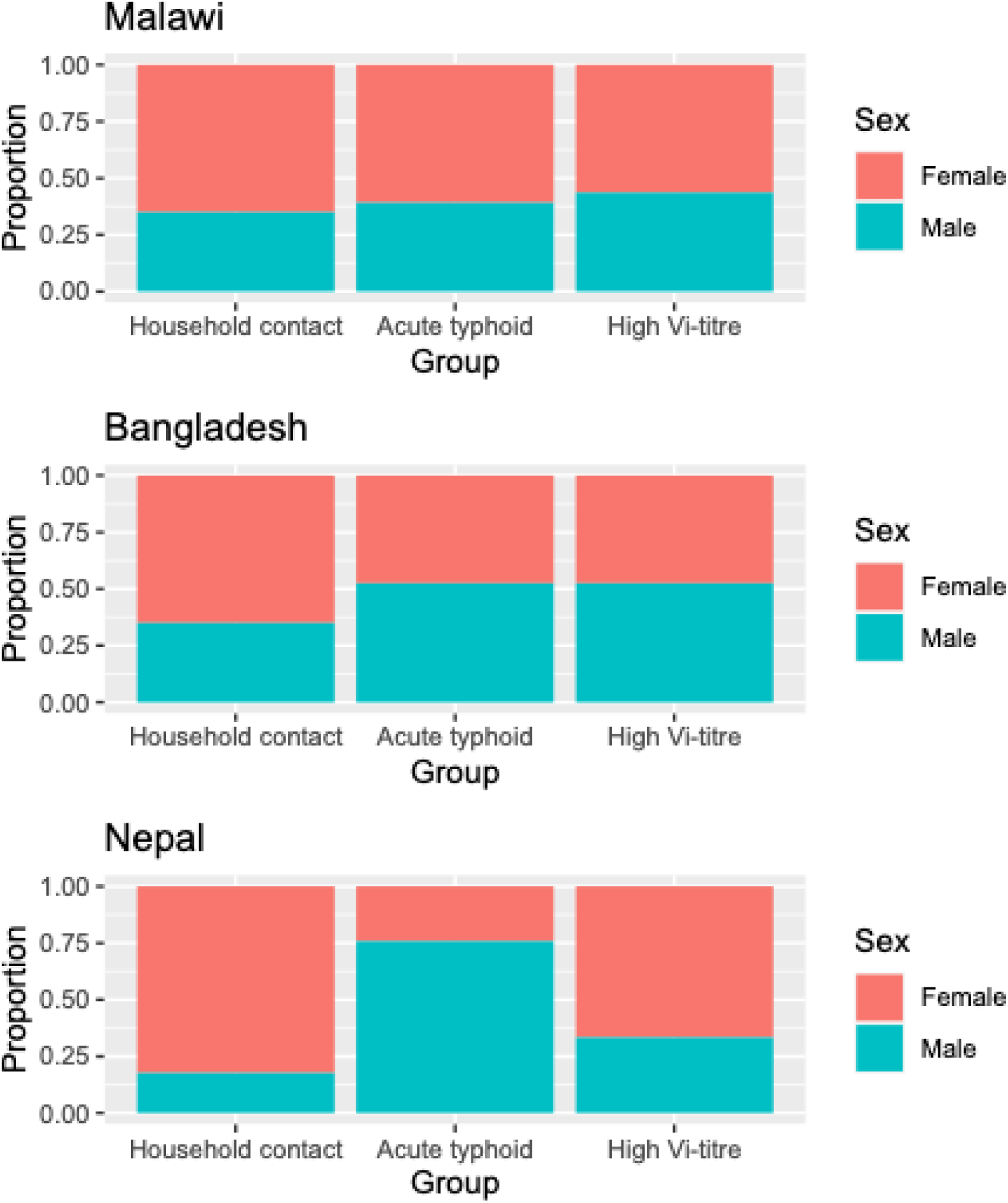
Sex split of the different participant groups across the different sites.

**Supplementary Figure 3:**
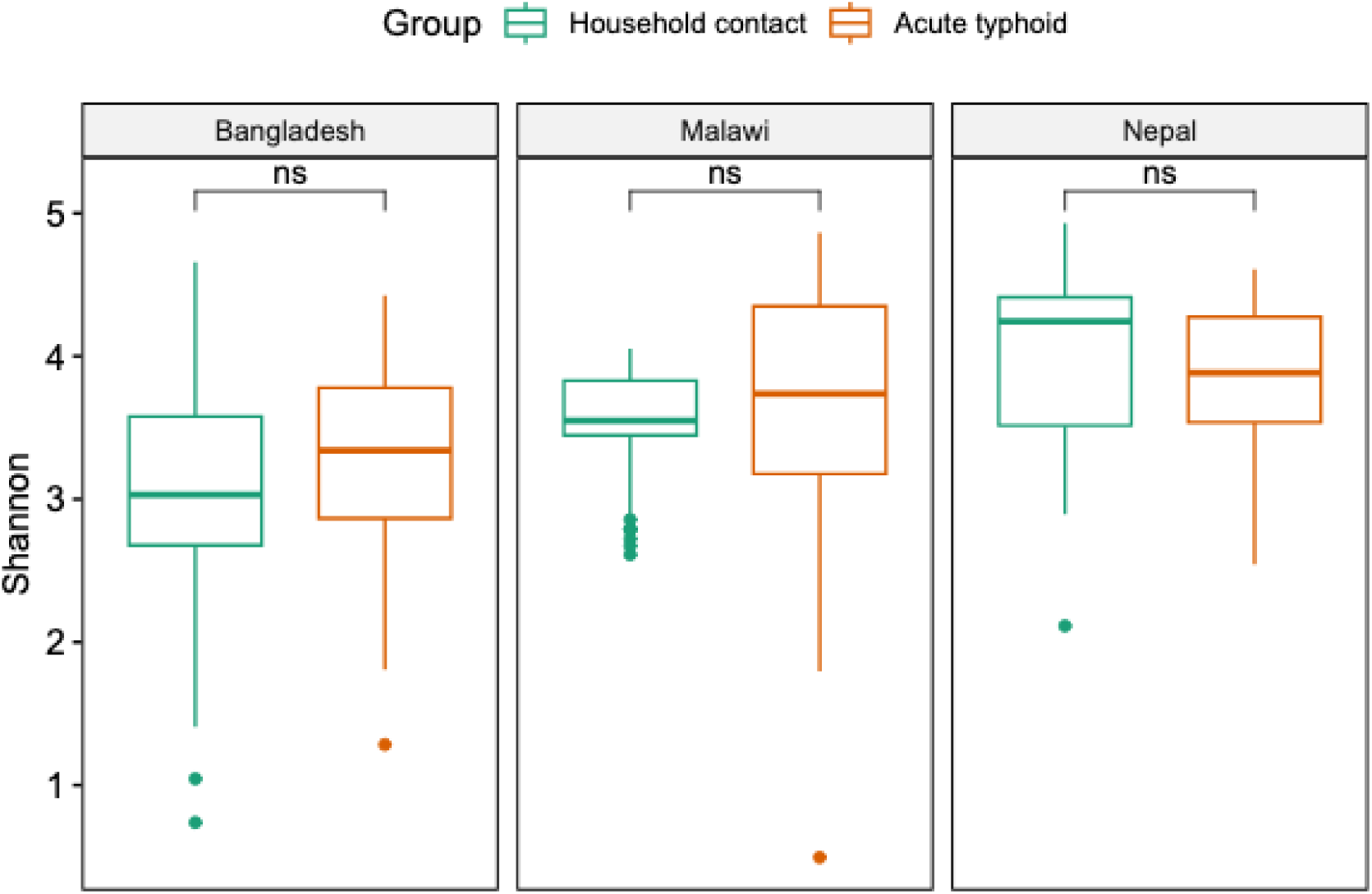
Alpha diversity for acute typhoid fever patients and healthy household contacts for each endemic country study site.

**Supplementary Figure 4:**
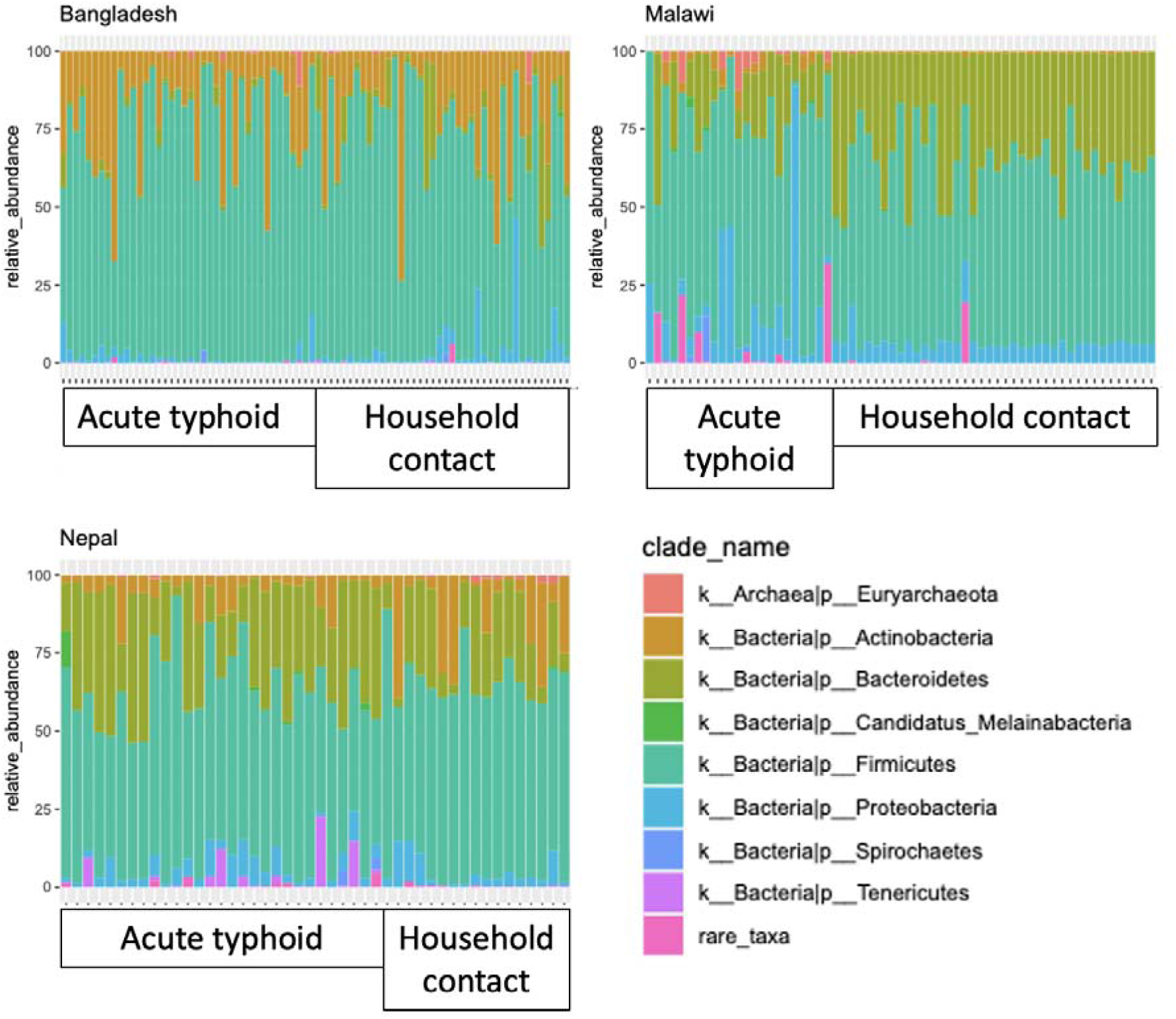
Bar chart showing the prevalence of eight common phyla in acute typhoid patients and healthy household contacts, for each study site.

**Supplementary Figure 5:**
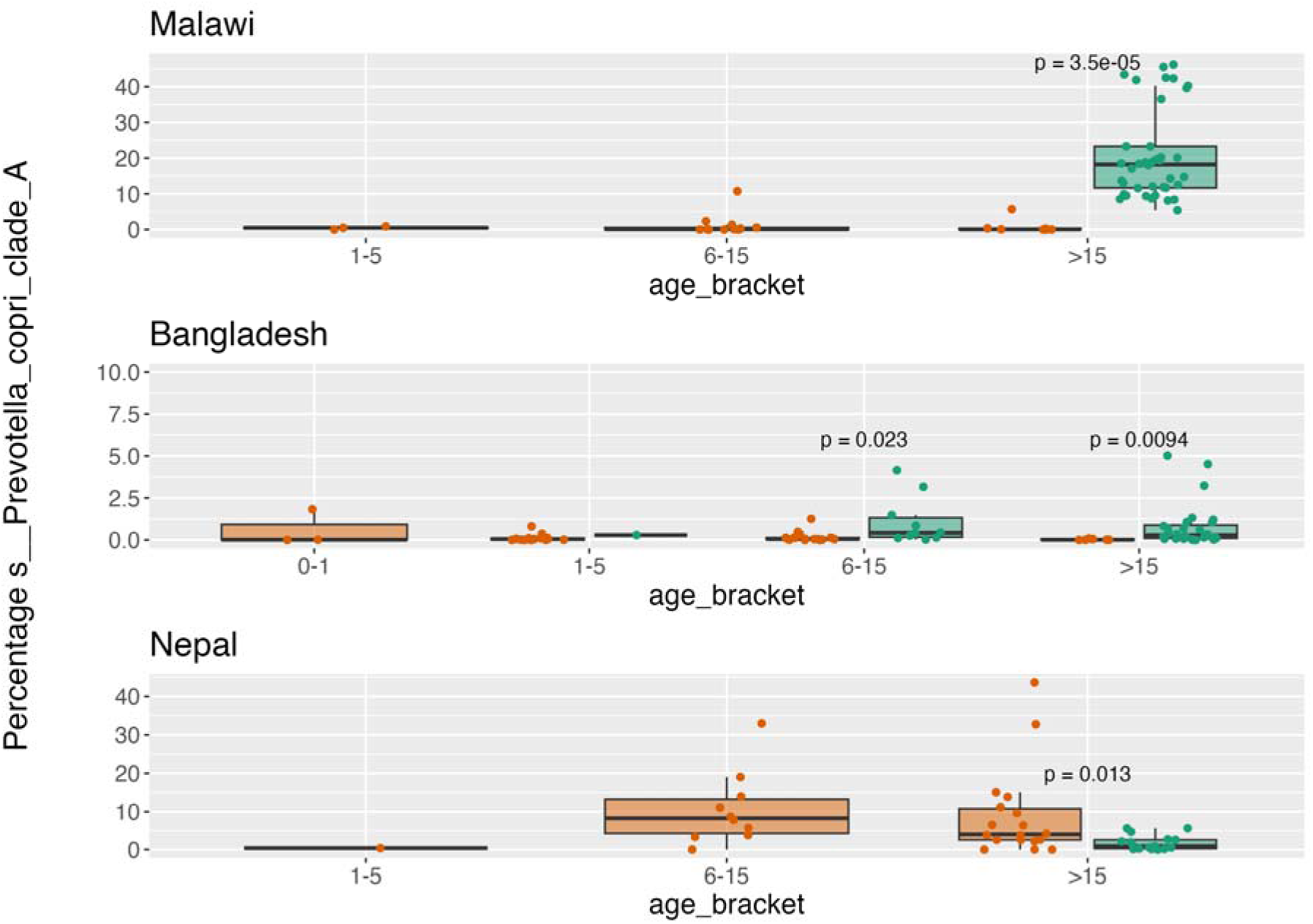
Prevalence of Prevotella copri clade A across age brackets in Malawi, Bangladesh, and Nepal.

**Supplementary Figure 6:**
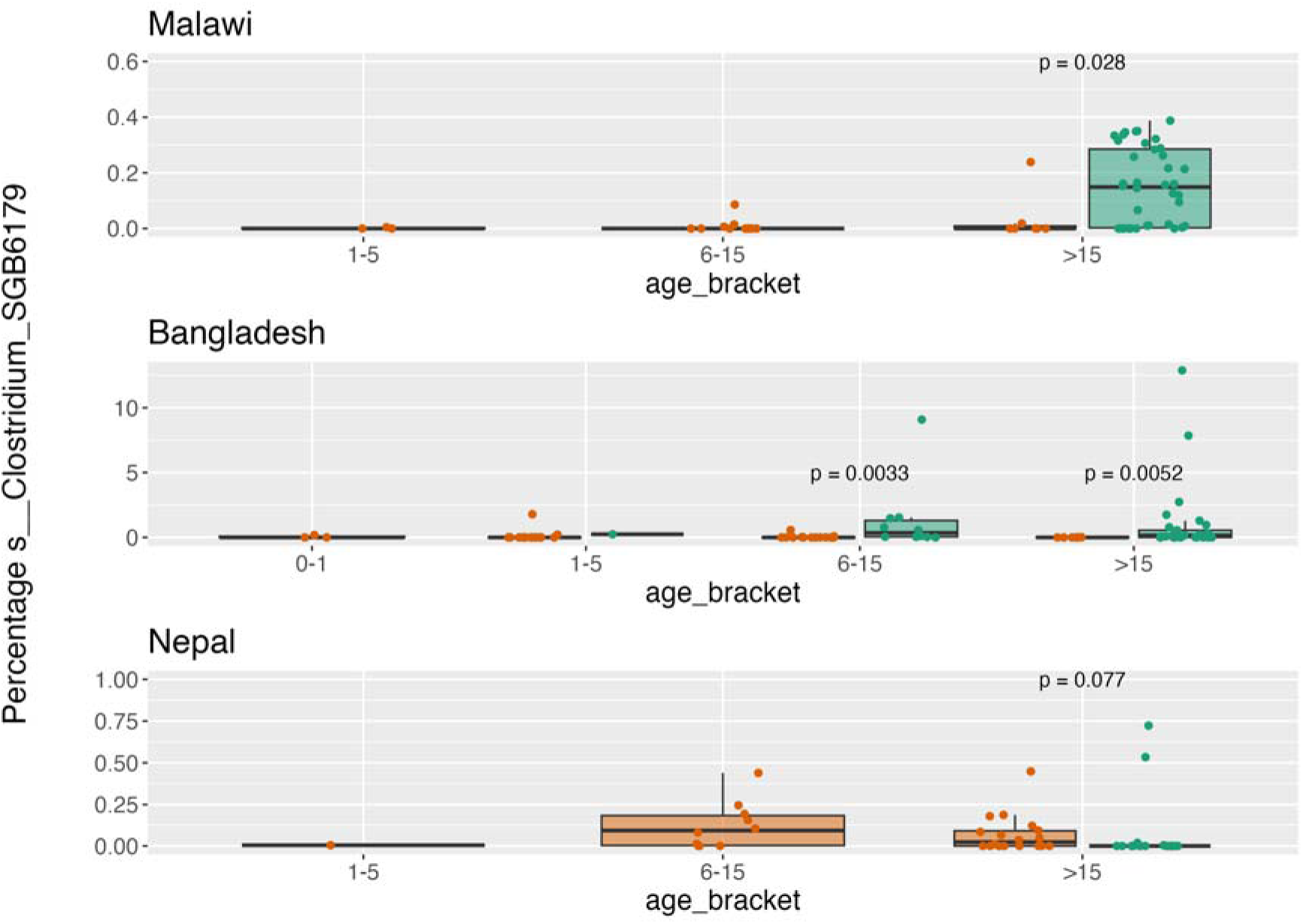
Prevalence of Clostridium SGB6179 across age brackets in Malawi, Bangladesh, and Nepal

**Supplementary Figure 7:**
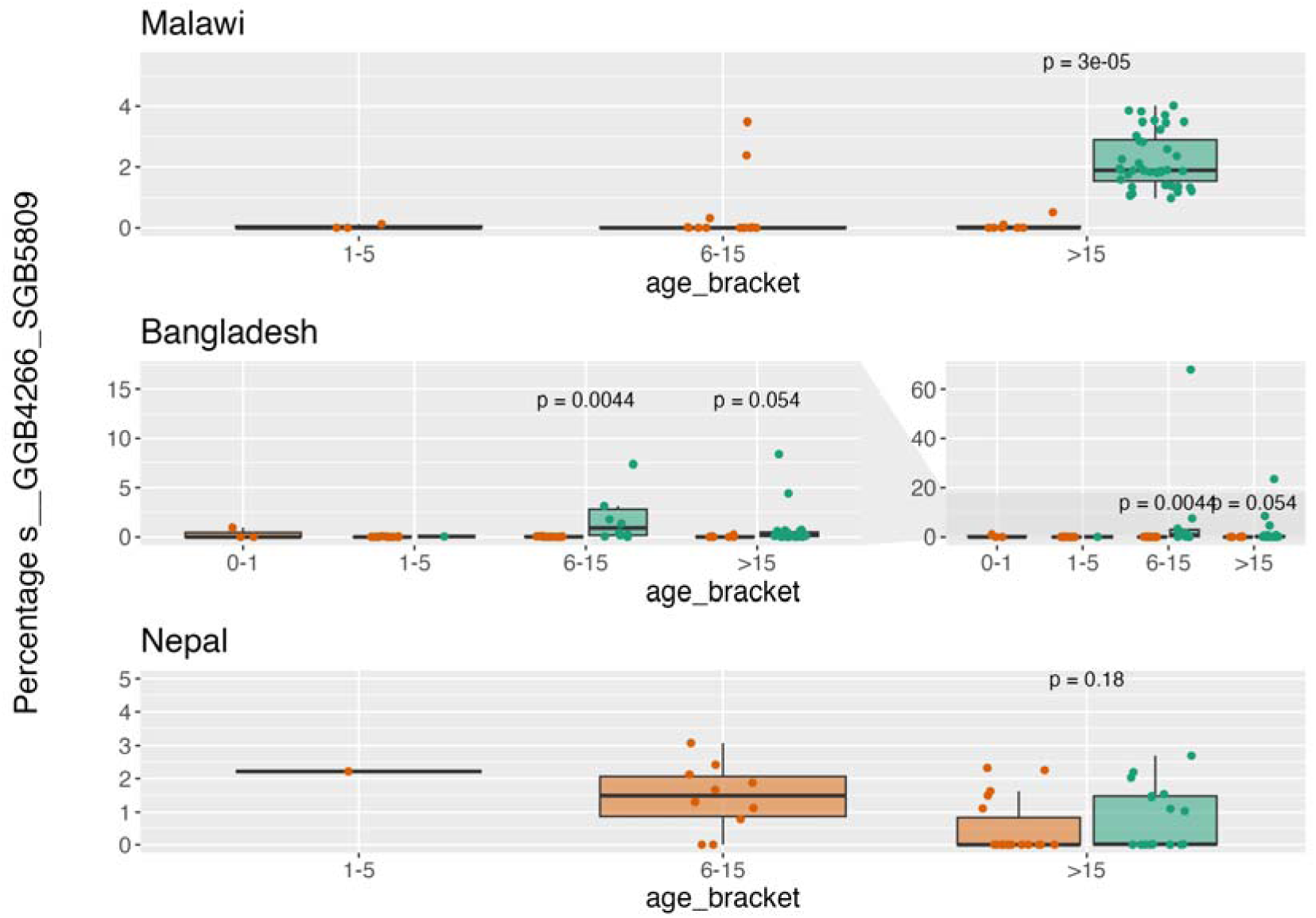
Prevalence of GGB4266_SGB5809 across age brackets in Malawi, Bangladesh, and Nepal

**Supplementary Figure 8:**
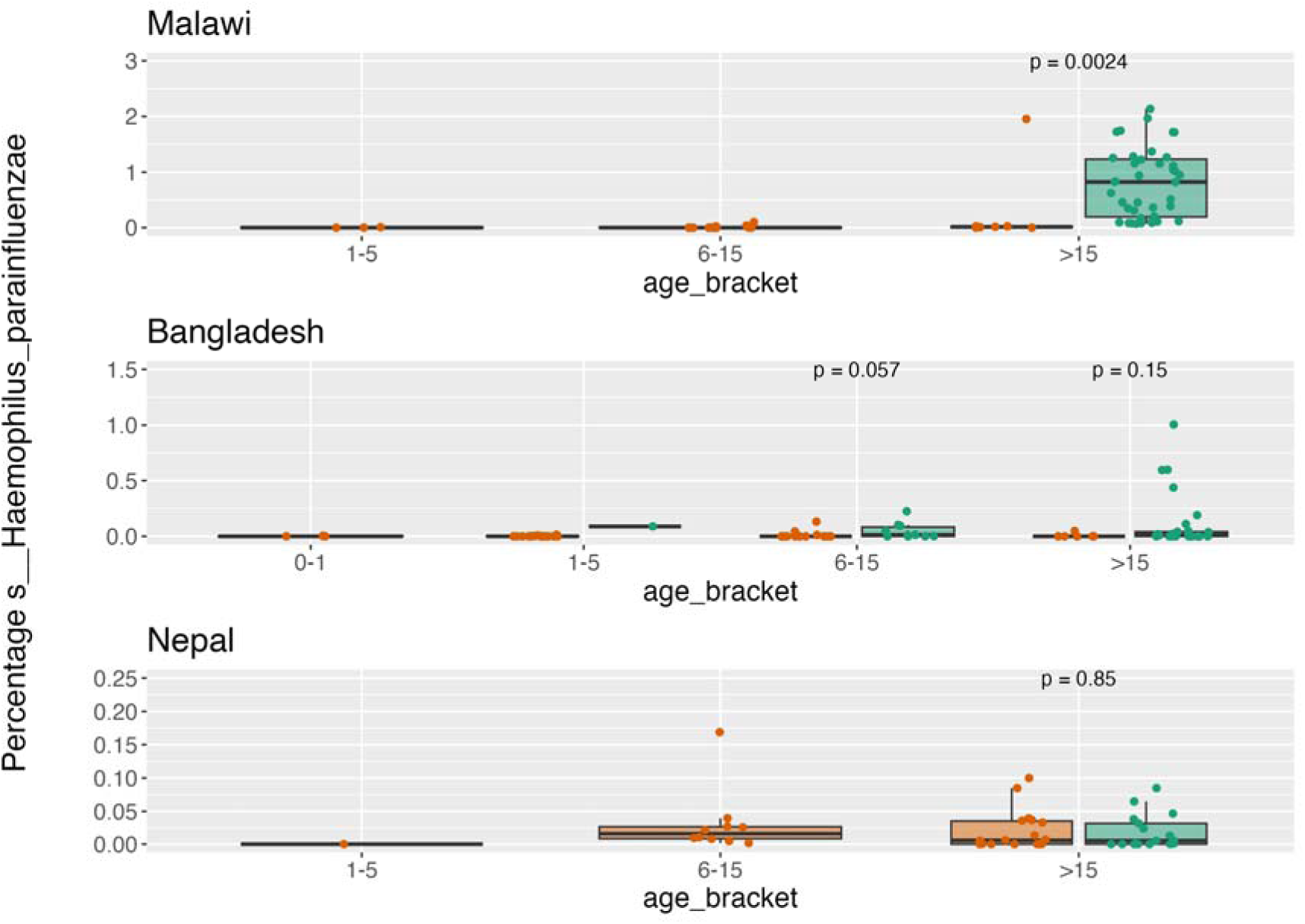
Prevalence of Haemophilus parainfluenzae across age brackets in Malawi, Bangladesh, and Nepal

**Supplementary Figure 9:**
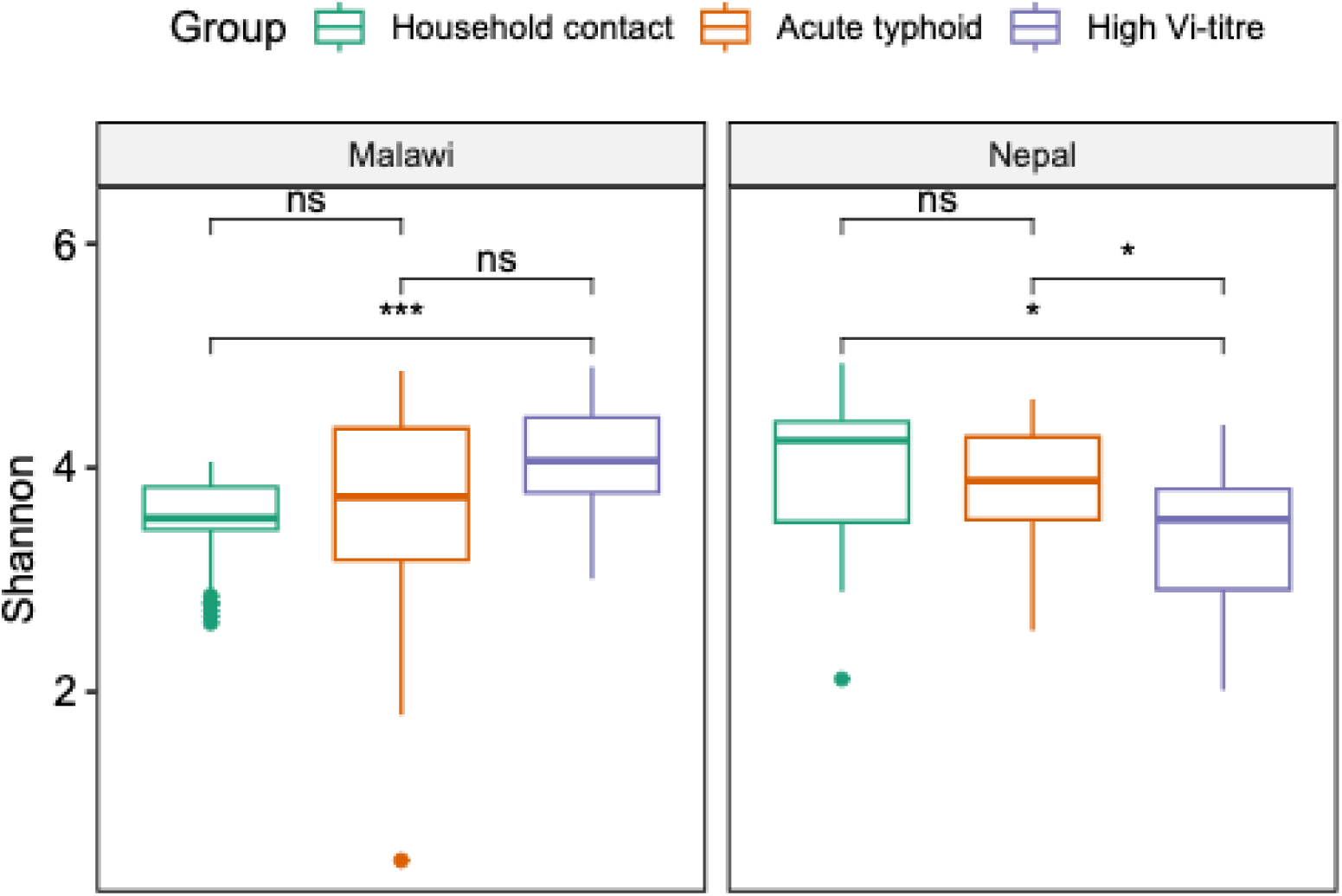
Alpha diversity of acute typhoid patients, high Vi-titre participants, and household contacts from Malawi and Nepal. Statistical annotations are the p-values resulting from Wilcoxon rank sum tests. A single asterisk indicates a p-value < 0.01, triple asterisk indicates a p-value < 0.0001.

**Supplementary Figure 10:**
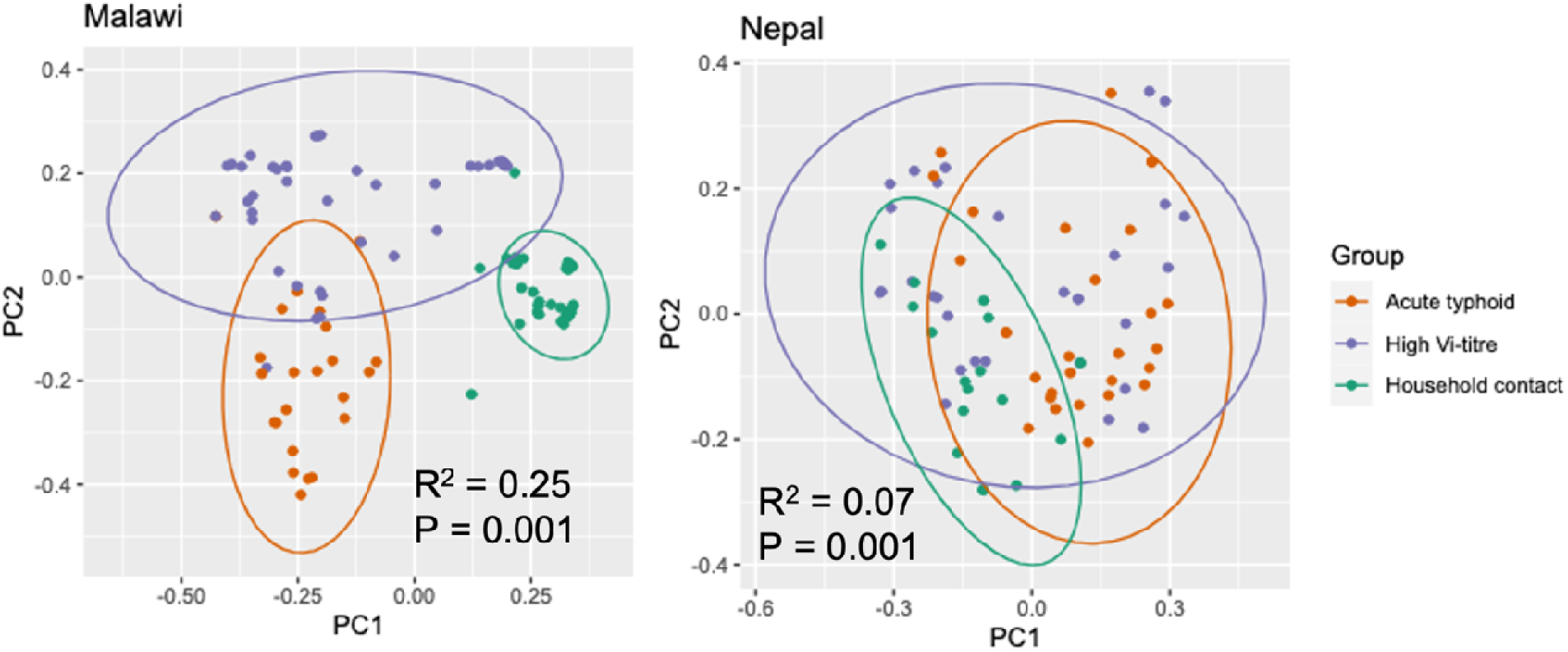
Beta diversity of acute typhoid fever, high Vi-titre participants, and household contacts from Malawi and Nepal

**Supplementary Figure 11:**
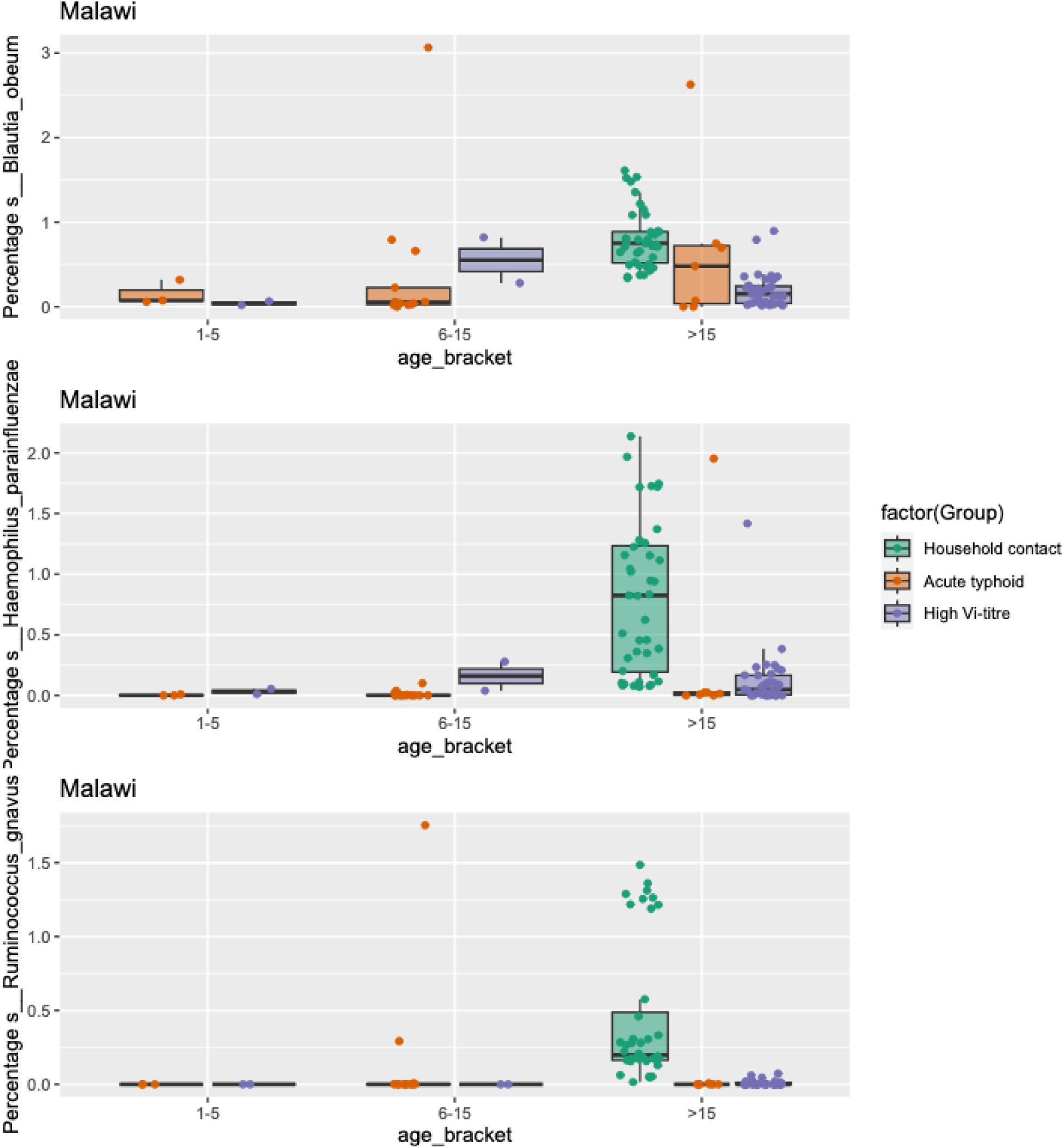
Prevalence of B. obeum, H. parainfluenzae, and R. gnavus across three participant groups (household contacts, typhoid cases, and high Vi-titre) in Malawi.

**Supplementary Figure 12:**
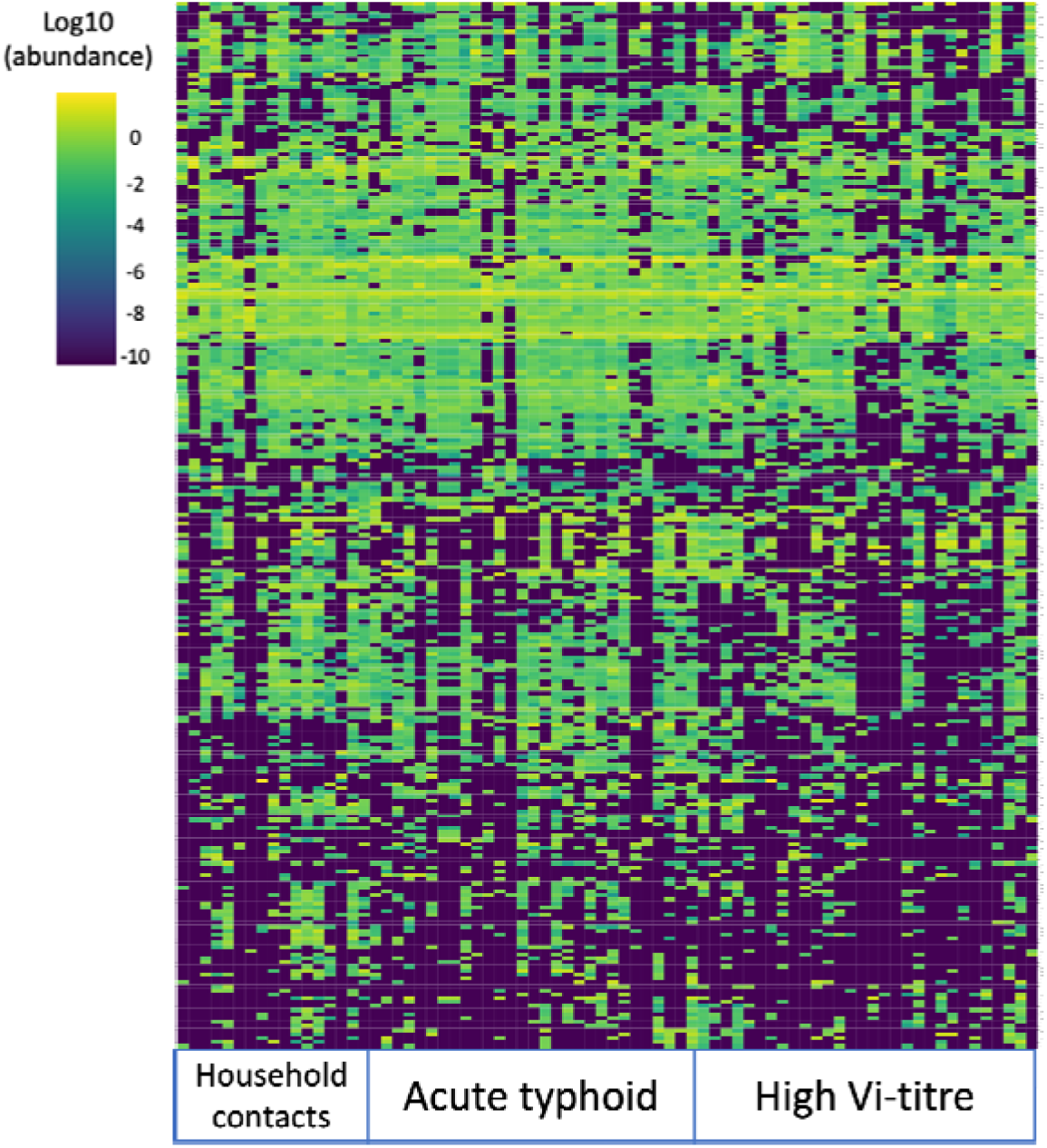
Heatmap of top 400 taxa from Nepalese participants – household contacts, acute typhoid cases and high-Vi titre participants.

**Supplementary Figure 13:**
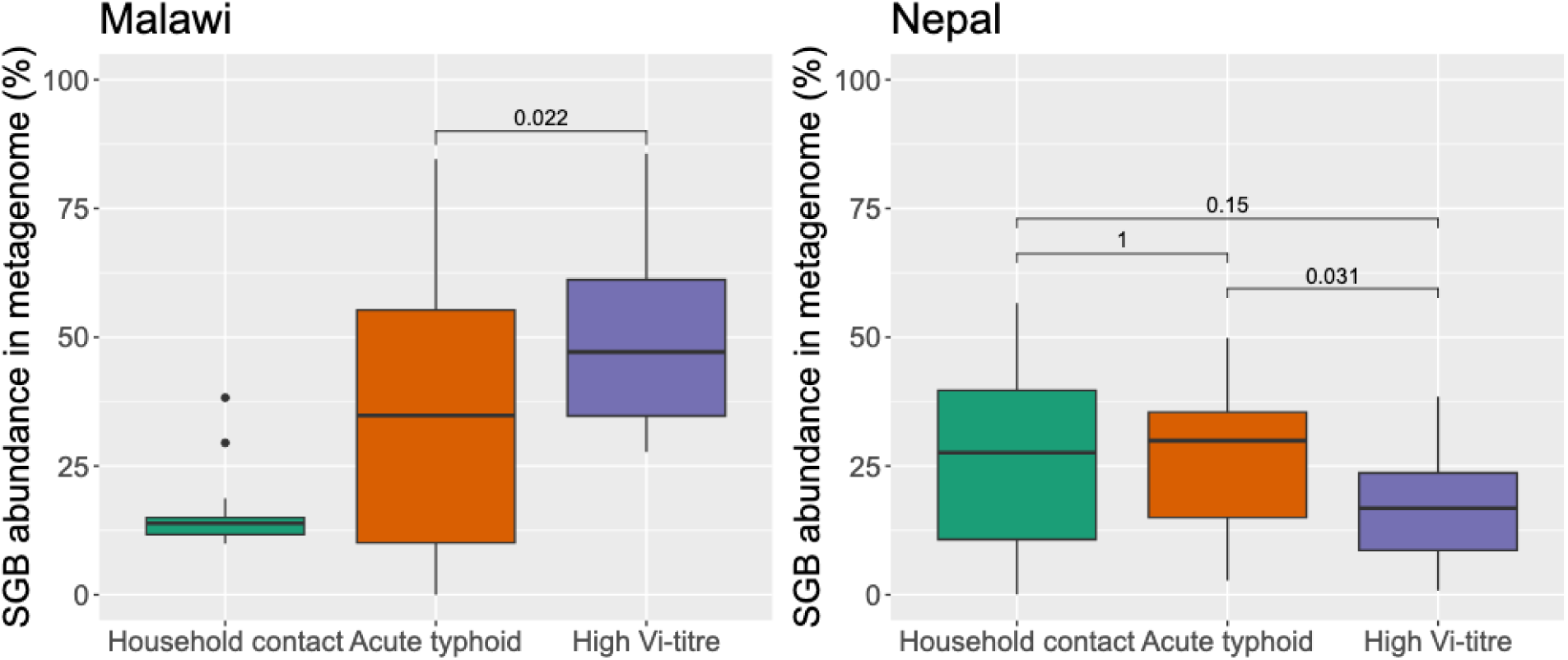
The abundance of species only described as species genome bins in the metaphlan database from Malawian and Nepalese household contacts, acute typhoid, and high-Vi titre participants.

**Supplementary Figure 14:**
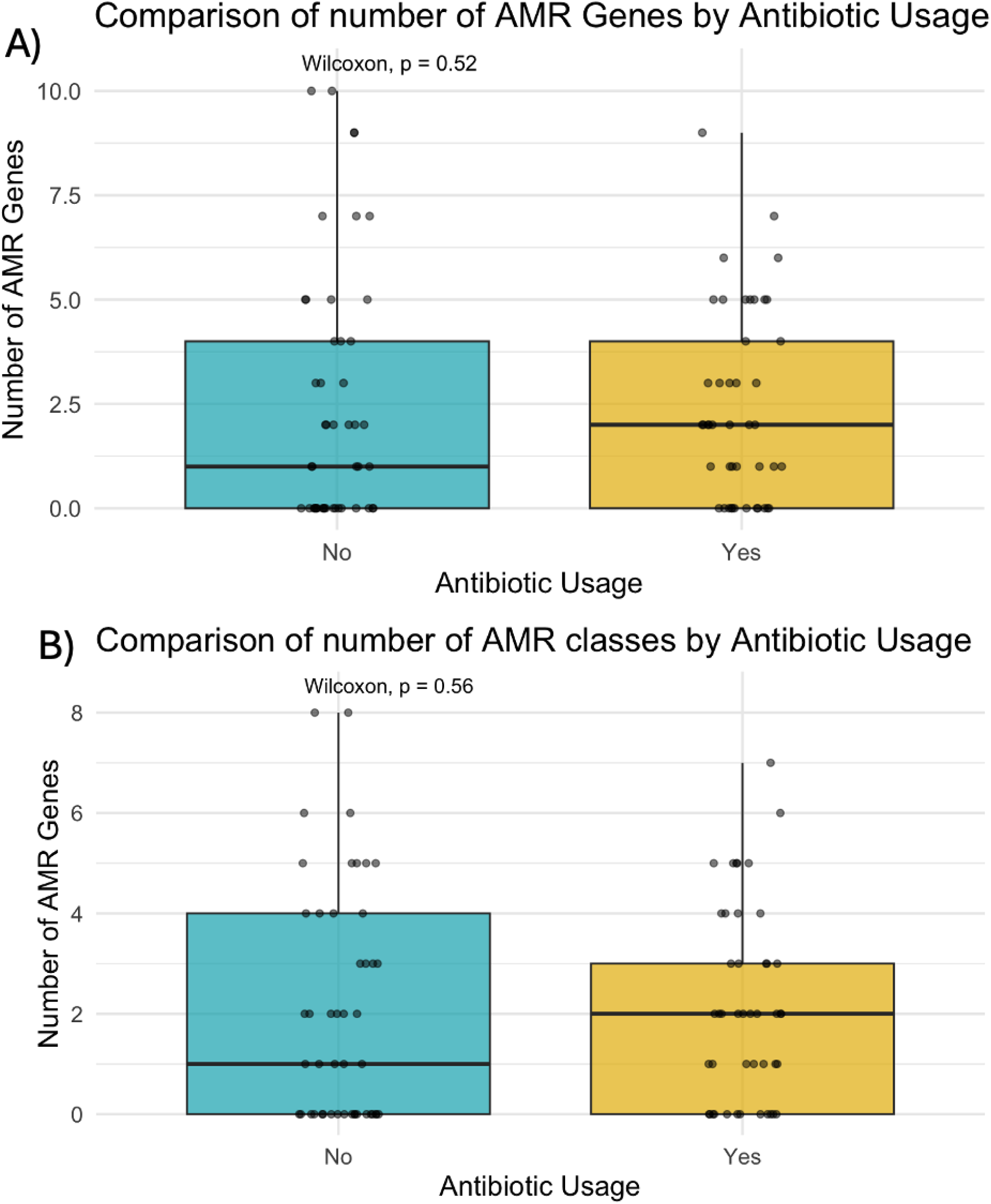
Comparison of A) the number of AMR gene and B) the number of AMR gene classes identified in participants from all three sites who did and did not report antibiotic usage prior to sampling

**Supplementary Figure 15:**
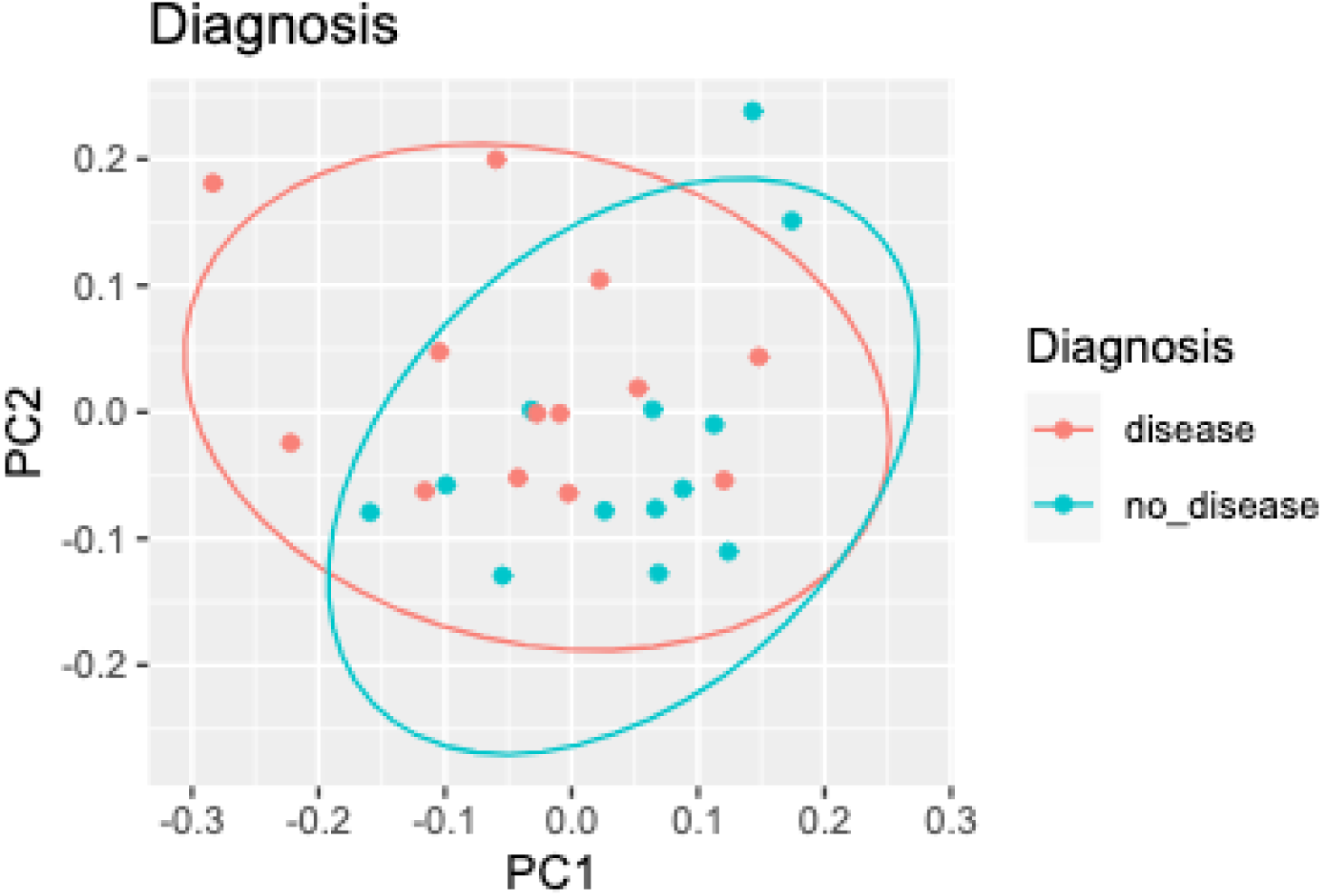
Beta-diversity plot of 26 CHIM participants challenged with S. Typhi or S. Paratyphi, coloured by whether they were diagnosed with typhoid/paratyphoid fever following pathogen exposure.

**Supplementary Figure 16:**
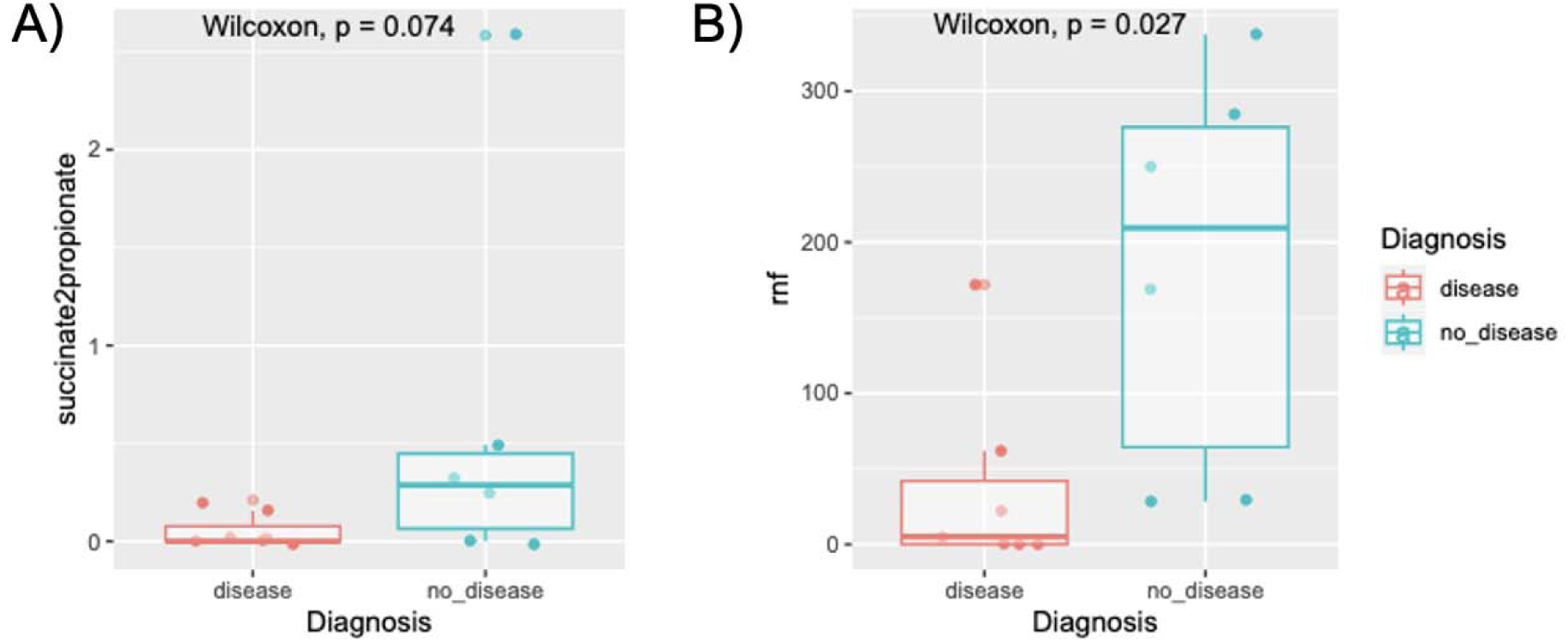
Comparison of prevalence of A) succinate2priopionate and B) rnf MGCs between people who were susceptible (disease) and non-susceptible (no_disease) to challenge with S. Typhi.

